# Identifying drivers of β-lactam/β-lactamase inhibitor resistance emergence and spread before their clinical deployment

**DOI:** 10.1101/2025.07.29.25331838

**Authors:** Kyle J Gontjes, Ali Pirani, Dhatri Badri Narayanan, Lisa J Lojek, Jennifer H Han, Pam Tolomeo, Ellie JC Goldstein, Ebbing Lautenbach, Evan Snitkin

## Abstract

Understanding the landscape of resistance to antibiotics before their clinical deployment could inform strategies to slow the development of resistance upon their introduction. We evaluated the associations between bacterial genotypes, patient clinical characteristics, and medical exposures with resistance to two recently approved β-lactam/β-lactamase inhibitor combinations in carbapenem-resistant *Klebsiella pneumoniae* collected before their clinical deployment. Whole-genome sequencing revealed that even within the clonal sequence type 258, genetic background influenced baseline resistance levels and the propensity for resistance to emerge and spread. Resistance in ST258 clade II was mediated by convergent mutation at a small number of loci, which rarely spread amongst patients. In contrast, resistance in clade I was influenced by a lineage-defining insertion in the OmpK36 porin, which made resistance accessible via the *bla*_KPC_ transposon moving to a higher copy number plasmid, and was in turn associated with transmissible resistance. Distinguishing patients based on whether resistance was acquired via plasmid-mediated versus mutational resistance revealed that while exposure to carbapenems and cephalosporins were associated with mutational resistance, plasmid-associated resistance in clade I was not associated with antibiotic exposures. These findings underscore how pre-clinical surveillance in clinically relevant populations can identify potential drivers of resistance that could inform molecular surveillance and antibiotic stewardship interventions to prolong the efficacy of novel antibiotics.

## INTRODUCTION

The emergence and spread of carbapenem-resistant *Klebsiella pneumoniae* (CRKP) poses a significant public health threat^1^. Of greatest concern are epidemic lineages that have acquired multidrug resistance, rendering standard therapies ineffective and increasing mortality rates^2^. The rising prevalence of multidrug-resistant CRKP, especially among critically ill patients, heightens the risk of poor outcomes in vulnerable populations^3^.

Several antibiotics were recently approved and introduced to treat these highly resistant infections^4^. Among the most notable therapeutics are β-lactam/β-lactamase inhibitor (BL/BLI) combinations, such as imipenem-relebactam (IR) and meropenem-vaborbactam (MVB)^5^. Despite promising initial reports of clinical effectiveness, concerns for their long-term viability have arisen due to reports of resistance in high-risk patient populations, even before these antibiotics reached the market^6–13^.

Our ability to limit the emergence and spread of resistance as antibiotics come to the market is hindered by an incomplete understanding of the bacterial and clinical factors that influence resistance emergence.

Previous reports have unsurprisingly demonstrated that antibiotic exposures exert selective pressure for resistance^14–17^. Whether other patient characteristics or exposures contribute to the emergence of antibiotic-resistant bacteria, particularly those with epidemic potential, is less clear. High rates of resistance emergence have been reported among patients with increased acuity^17,18^. In addition, variation in rates of resistance emergence across healthcare settings have also been reported, but this could be due to differences in case-mix, transmission risk, or antibiotic stewardship and infection prevention practices^19,20^.

Even less is understood about how bacterial genetic background potentiates the emergence and spread of antibiotic resistance. Support for the role of genetic background comes from observed lineage-specific differences in the prevalence and fitness cost of resistance-conferring genotypes and plasmids^21,22^. Further support comes from strictly *in vitro* studies, which have shown that chromosomal and plasmid backgrounds can affect strain fitness and transcriptional responses upon the acquisition of resistance-conferring plasmids^23–25^. Pre-existing chromosomal mutations have also been postulated to influence not just fitness costs, but also potentiate the rate of antibiotic resistance emergence^26–28^. Understanding how bacterial genetic background and patient clinical characteristics jointly influence resistance emergence and spread could strengthen our ability to predict and prevent resistance development.

To better understand how resistance to BL/BLI combinations emerges and spreads, we conducted a genomic epidemiology investigation using a comprehensive set of CRKP clinical isolates collected from a long-term acute care hospital (LTACH) network between August 2014 and July 2015, several years before the clinical deployment of MVB and IR. Genomic analysis provided evidence of repeated emergence and spread of resistant strains belonging to the epidemic lineage sequence type 258 (ST258). Genome-wide association studies (GWAS) and phylogenetic analysis demonstrated that the genetic background within ST258 influenced baseline resistance levels, the likelihood of resistance crossing clinical breakpoints, and the spread of resistant strains. Finally, risk factor analysis for patients whose resistant isolate was predicted to be acquired via carbapenemase or non-carbapenemase mechanisms uncovered that patient characteristics and antibiotic exposures may differentially contribute to these distinct routes to BL/BLI resistance. Together, these findings highlight the importance of pre-clinical surveillance in identifying both bacterial genetic and clinical drivers of resistance, which can inform molecular surveillance and interventions to prolong the efficacy of novel antibiotics.

## RESULTS

### Detection of resistance to β-lactam/β-lactamase inhibitor combinations before their clinical deployment

We collected 412 CRKP ST258 isolates from August 2014 to July 2015 across 21 LTACHs, including 232 respiratory, 139 urinary, 35 blood, and 6 wound isolates. When performing susceptibility testing for several last-line and novel antibiotics^6^, we observed that the median minimum inhibitory concentration (MIC) was 0.50 µg/mL (log_2_ interquartile range [IQR], 1.0) for IR and 0.25 µg/mL (log_2_ IQR, 2.0) for MVB, both in the susceptible range according to 2021 Clinical & Laboratory Standards Institute breakpoints (**Figure 1**). However, when comparing strict susceptibility, 58 (14.1%) isolates were resistant to at least one of the BL/BLI combinations: 39 (9.5%) to MVB and 38 (9.2%) to IR. Due to cross-resistance and a strong positive Spearman’s correlation between MICs (**Supplementary** Figures 1-2), downstream analyses evaluated BL/BLI resistance as a composite outcome.

**Figure 1.**
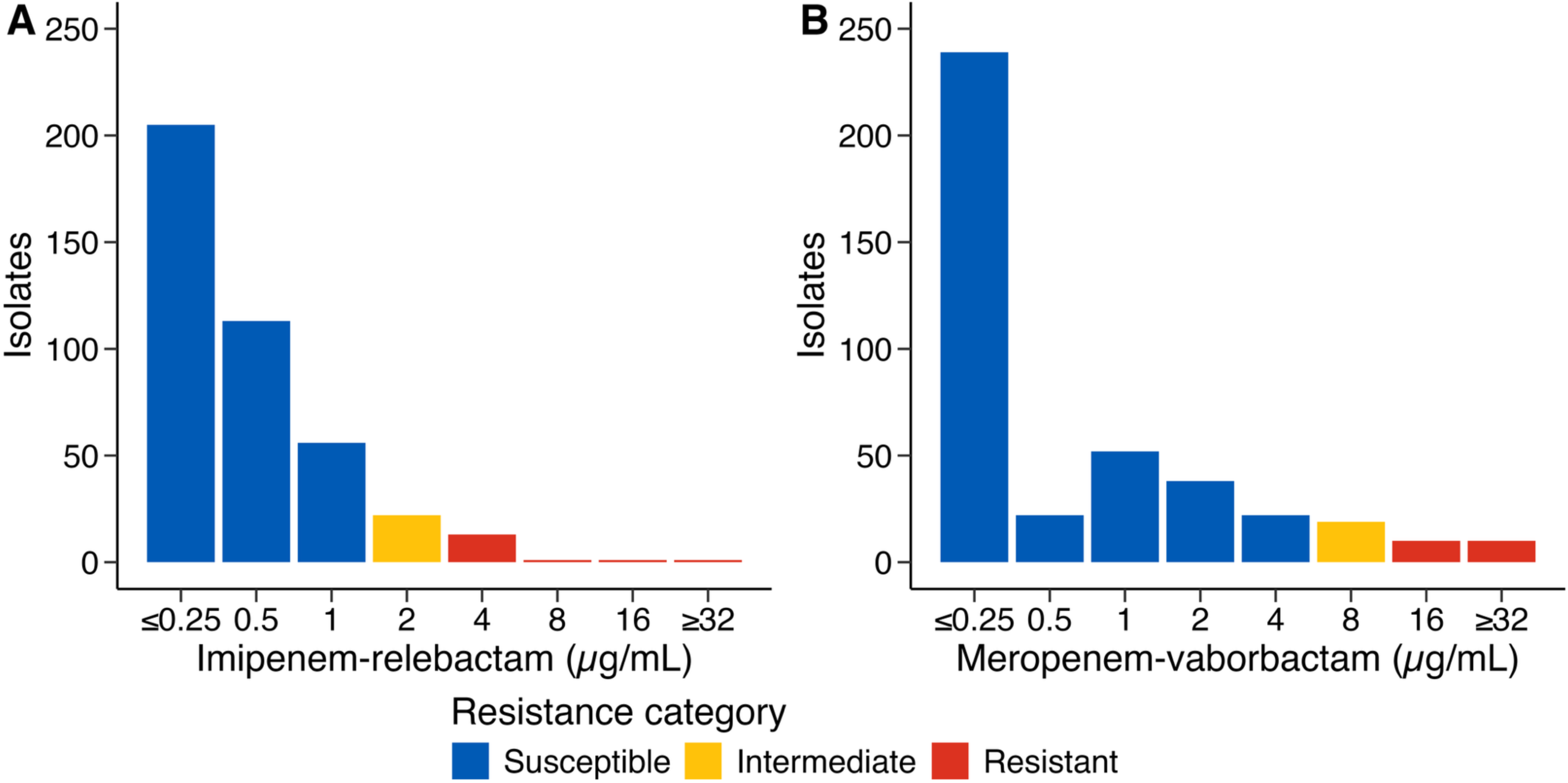
Minimum inhibitory concentrations for imipenem-relebactam and meropenem-vaborbactam in carbapenem-resistant *Klebsiella pneumoniae* ST258 isolates Distribution of minimum inhibitory concentrations for (**a**) imipenem-relebactam and (**b**) meropenem-vaborbactam, as measured using a custom broth microdilution panel (SENSITITRE, Thermo Fisher Scientific). The 2021 Clinical and Laboratory Standards Institute breakpoints were used to categorize the susceptibility profiles of these isolates. The β-lactamase inhibitor concentrations were 4 µg/mL and 8 µg/mL for imipenem-relebactam and meropenem-vaborbactam, respectively.

### Differences in resistance emergence and spread exist across clades of epidemic-lineage ST258

As previously reported^29,30^, our ST258 phylogeny had two clades: clade I (145 isolates [35.2%]) and clade II (267 isolates [64.8%]). Overlaying resistance phenotypes on the phylogeny revealed striking differences in resistance between the two clades (**Figure 2**). First, baseline MICs were significantly elevated in clade I for both IR and MVB (Wilcoxon rank-sum test p-values <0.001). Moreover, resistance, with MICs above clinical breakpoints, was significantly more frequent in clade I (39 [26.9%]) than in clade II (19 [7.1%]) (X^2^ test p-value <0.001).

**Figure 2.**
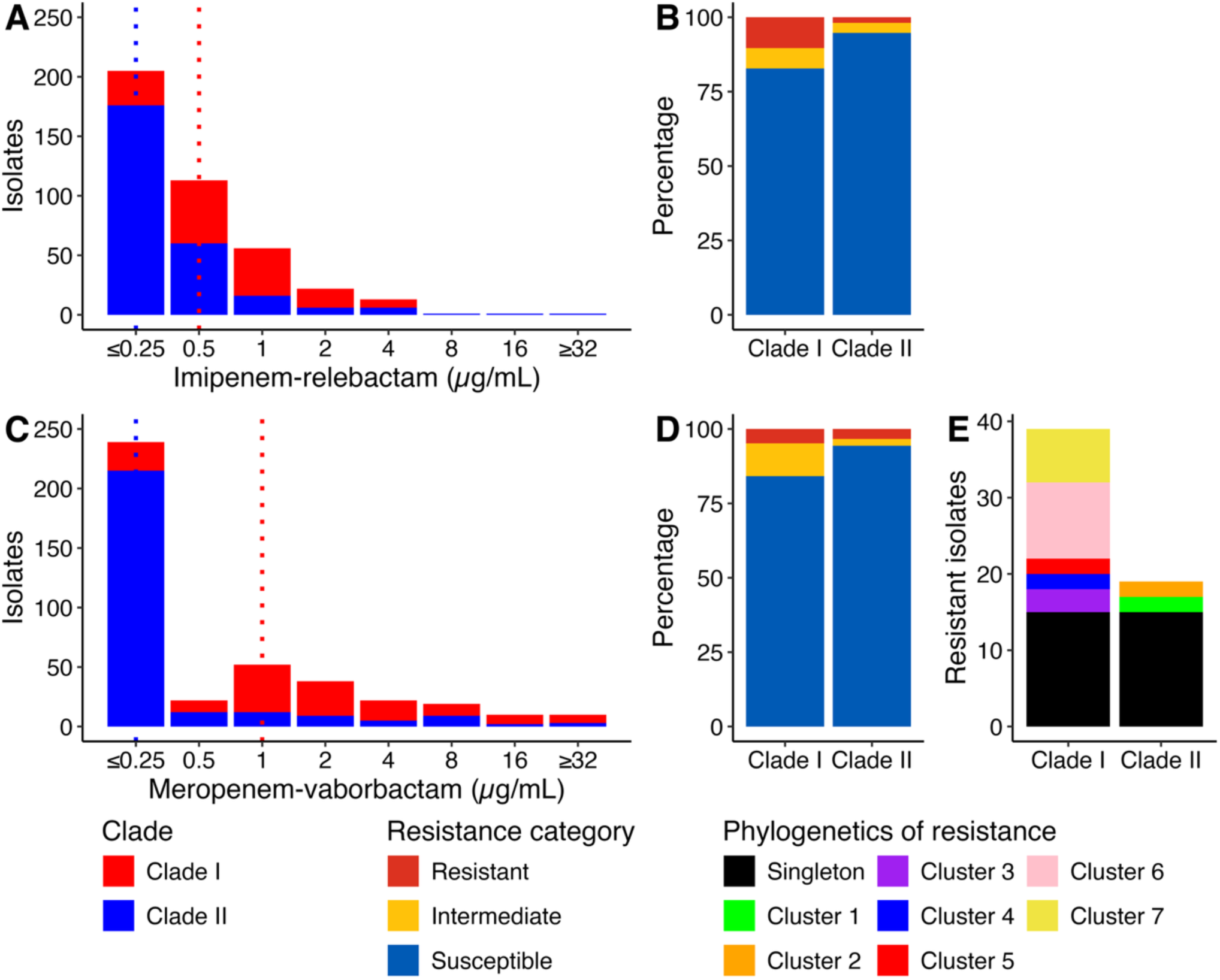
Differences in β-lactam/β-lactamase inhibitor combination resistance across clades of epidemic lineage sequence type 258 Minimum inhibitory concentration (MIC) distributions stratified by clades of sequence type 258 for (**a**) imipenem-relebactam and (**c**) meropenem-vaborbactam. Dashed vertical lines indicate the median MIC values for each clade. The 2021 Clinical and Laboratory Standards Institute breakpoints were used to categorize the susceptibility profile for (**b**) imipenem-relebactam and (**d**) meropenem-vaborbactam. The membership of resistant isolates in phylogenetic clusters and singletons (**e**).

To better understand the historical basis for elevated resistance in clade I, ancestral state reconstruction of BL/BLI resistance was performed to identify independent resistance emergence events. Using this approach, the 58 resistant isolates were partitioned into 36 emergence events. Twenty-nine emergence events involved only one patient, indicating repeated *de novo* evolution of resistance without subsequent spread. Of note, these 29 emergence events were split between the two clades, with 15 in clade I and 14 in clade II, despite there being almost twice as many clade II isolates were collected. Consistent with the increased frequency of resistance emergence in clade I observed with ancestral reconstruction, we detected significant differences in the phylogenetic rates of resistance emergence (likelihood ratio test p-value < 0.001), with higher rates of phenotypic transition in clade I (q_transition_, 20965.15) than in clade II (q_transition_, 3881.492). In addition to resistant singletons, seven phylogenetic clusters of resistance involving multiple patients were detected, consistent with the emergence and spread of resistant strains: five in clade I and two in clade II. Although not statistically significant (Fisher’s exact test p-value = 0.43), the proportion of emergence events that spread among patients was also elevated in clade I (5 clusters / 20 events [25.0%]), as compared to clade II (2 clusters / 16 events [12.5%]). These results suggest that ST258 clade I had an increased propensity for both the evolution and cross-transmission of BL/BLI resistance, supporting a role for bacterial genetic background in the dynamics of resistance to these therapeutics.

### Non-carbapenemase mechanisms of carbapenem resistance explain BL/BLI resistance in a clade-specific manner

We next sought to understand the genetic basis for BL/BLI resistance across the two ST258 clades. As both β-lactamase inhibitors inhibit the *bla*_KPC_ carbapenemase ubiquitous in ST258 (408/412 [99.0%]), we hypothesized that resistance was driven by non-carbapenemase mechanisms of carbapenem resistance^5,31–33^. To test this hypothesis, genotypes involved in carbapenem entry, efflux, and target modification were overlaid on the phylogeny in **Supplementary** Figure 3, and their frequency in resistant isolates is reported in **Table 1**.

**Table 1.** Presence of non-carbapenemase mechanisms of carbapenem resistance stratified by resistance to β-lactam/β-lactamase inhibitor combinations.

| Mechanism of interest | Overall<br>(n=412) | BL/BLI Resistant<br>(n=58) | BL/BLI Susceptible<br>(n=354) | p-value | Nearest-neighbor<br>criterion | Discriminatory<br>criterion |
| --- | --- | --- | --- | --- | --- | --- |
| **Carbapenem entry** |  |  |  |  |  |  |
| *ompK35* porin truncation at 25% | 412 (100.0) | 58 (100.0) | 354 (100.0) | NA | NA | NA |
| *ompK36* porin | 131 (31.8) | 37 (63.8) | 94 (26.6) | <0.001 | Pass | Fail |
| Cytosine-to-thymine transition at position 25 | 124 (30.1) | 37 (63.8) | 87 (24.6) | <0.001 | Pass | Fail |
| Loop 3 insertion | 119 (28.9) | 35 (60.3) | 84 (23.7) | <0.001 | Pass | Fail |
| GD insertion | 5 (1.2) | 2 (3.4) | 3 (0.8) | 0.147 | Fail | Pass |
| Non-synonymous mutation | 22 (5.3) | 10 (17.2) | 12 (3.4) | <0.001 | Pass | Pass |
| Putative function-altering variants | 20 (4.9) | 10 (17.2) | 10 (2.8) | <0.001 | Pass | Pass |
| Truncation | 8 (1.9) | 6 (10.3) | 2 (0.6) | <0.001 | Pass | Pass |
| *ompK36* 5' intergenic region |  |  |  |  |  |  |
| Any mutation | 31 (7.5) | 8 (13.8) | 23 (6.5) | 0.061 | Fail | Pass |
| Insertion sequences | 24 (5.8) | 6 (10.3) | 18 (5.1) | 0.128 | Pass | Pass |
| **Carbapenem efflux** |  |  |  |  |  |  |
| *acrAB-tolC* efflux pump mutant | 2 (0.5) | 0 (0.0) | 2 (0.6) | 1.000 | Fail | Pass |
| *ramR* efflux pump regulator putative-function altering variants | 27 (6.6) | 7 (12.1) | 20 (5.6) | 0.083 | Fail | Pass |
| *ramA* efflux pump activator putative-function altering variants | 10 (2.4) | 6 (10.3) | 4 (1.1) | 0.001 | Pass | Pass |
| **Carbapenem target modification** |  |  |  |  |  |  |
| Non-synonymous mutations in penicillin-binding-proteins | 9 (2.2) | 6 (10.3) | 3 (0.8) | <0.001 | Pass | Pass |
| Penicillin-binding protein-2 | 8 (1.9) | 6 (10.3) | 2 (0.6) | <0.001 | Pass | Pass |
| Penicillin-binding protein-4 | 2 (0.5) | 1 (1.7) | 1 (0.3) | 0.262 | Pass | Pass |

The frequency of non-carbapenemase mechanisms of carbapenem resistance was stratified by resistance to at least one β-lactam/β-lactamase inhibitors (BL/BLI) combination: imipenem-relebactam and meropenem-vaborbactam. Statistical significance was evaluated using Fisher’s exact test due to a small number of events, except for the presence of any loop 3 insertion or the GD insertion in loop 3 of *ompK36*, where the X<sup>2</sup> test was performed. To account for confounding by genetic background, the MICs of isolates with the genotype were compared to their nearest phylogenetic neighbor without the genotype. Genotypes with a median log2 fold change in MIC > 1 passed our nearest neighbor criterion. Genotypes with discriminatory potential (92.5% specificity and 80% accuracy) passed the discriminatory potential criterion. **Abbreviations:** BL/BLI, β-lactam/β-lactamase inhibitor; NA, not applicable.

To account for confounding by genetic background and identify associations more likely to be causal, discriminatory genotypes were identified by comparing the MICs of isolates with each putative resistance genotype to their nearest phylogenetic neighbor without the genotype (**Supplementary** Figure 3C). The following genotypes had discriminatory potential (92.5%+ specificity and 80% accuracy) and a median log_2_ fold change in MIC > 1, rendering them eligible for inclusion into a panel of explanatory genotypes: mutations in *ompK36* (truncations and putative function-altering variants), insertion sequences near the *ompK36* promoter, putative function-altering variants in *ramA*, and non-synonymous mutations in penicillin-binding protein (PBP) genes. Genotypes passing these criteria were identified in 22 of 58 (37.9%) BL/BLI-resistant isolates. Notably, there were marked differences in both the frequency and explanatory power of carbapenem resistance-associated genotypes across clades (**Supplementary Table 1; Supplementary** Figure 3). Resistance in clade II was almost completely accounted for by variation in this curated gene set (15/19 [78.9%]), whereas resistance in clade I was left largely unexplained (7/39 [17.9%]).

### Convergence-based genome-wide association studies improved the explanation of resistance and illuminated distinct pathways to resistance across clades

As most resistant isolates were not explained by evaluated non-carbapenemase mechanisms of carbapenem resistance, we next sought to identify alternative genetic mechanisms. To identify novel determinants of resistance, convergence-based GWAS were performed on continuous log_2_ MIC and discrete BL/BLI resistance. GWAS performed on continuous MIC values sought to identify modulators of resistance, while GWAS performed on discrete resistance categorization sought determinants of transitions across the clinical breakpoint. Ten significant hits were detected (**Supplementary** Figure 4), eight of which passed the nearest neighbor criterion and were considered further (four continuous and four discrete, **Figure 3**).

**Figure 3.**
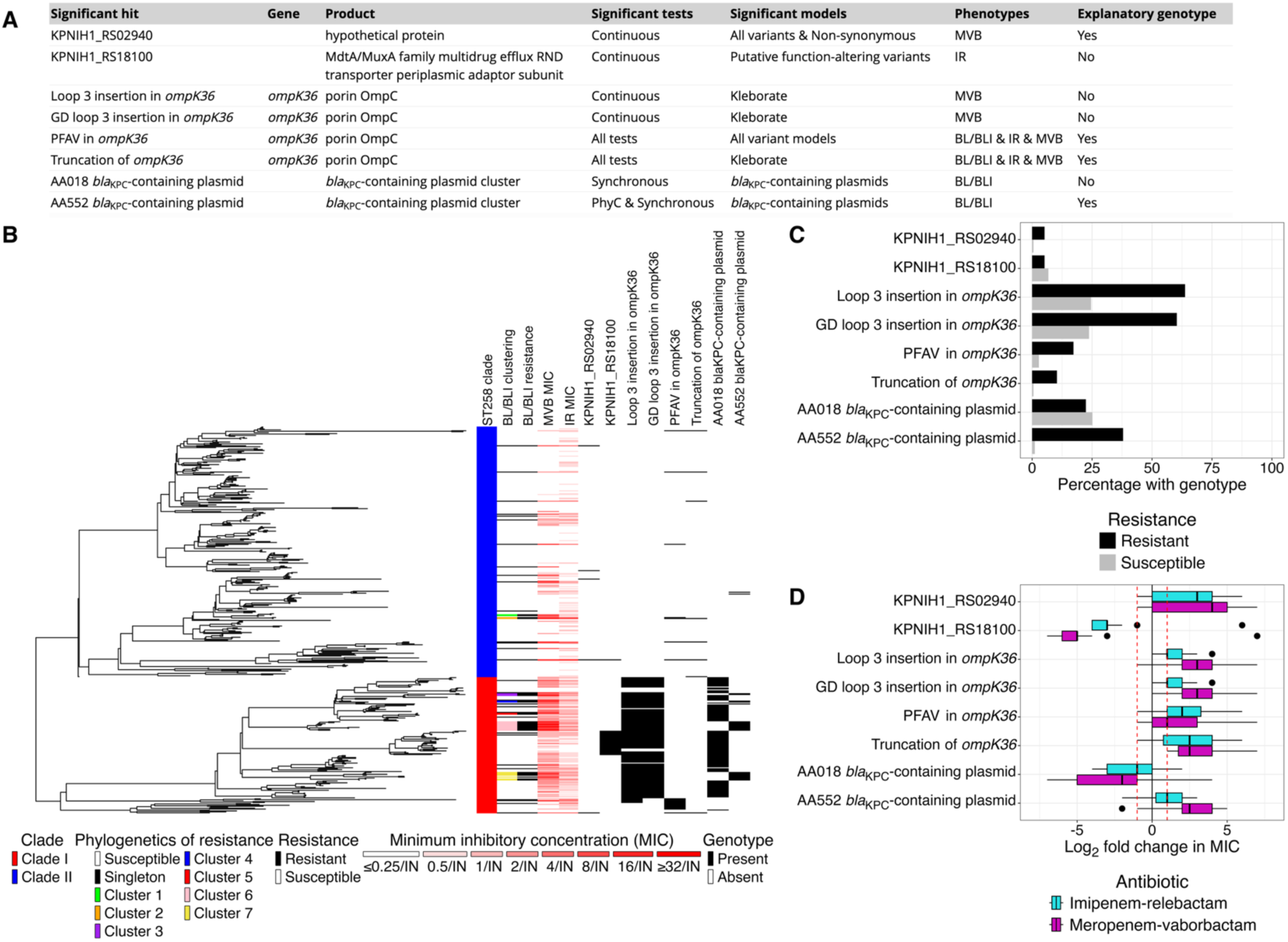
Qualifying genome-wide association study hits for β-lactam/β-lactamase inhibitor resistance Convergence-based genome-wide association study was performed on log_2_ minimum inhibitory concentration (MIC) and resistance to β-lactam/β-lactamase inhibitor (BL/BLI) combinations using hogwash v1.2.6. Hits that passed the nearest neighbor criterion (median log_2_ fold change in MIC > |1|) were eligible for downstream analysis and included in this figure. (**a**) Information about each significant genome-wide association study hit. Genotypes were classified as explanatory if their median log_2_ fold change in MIC > 1 and had diagnostic potential (specificity > 92.5% and accuracy >80%). (**b**) The significant hits are overlaid on the phylogeny. (**c**) The frequency of genotypes in β-lactam/β-lactamase inhibitor combination susceptible and resistant isolates. (**d**) The log_2_ fold change in imipenem-relebactam and meropenem-vaborbactam minimum inhibitory concentration for isolates with the genotype relative to their nearest phylogenetic neighbor without the genotype. **Abbreviations:** BL/BLI, β-lactam/β-lactamase inhibitor; IR, imipenem-relebactam; MVB, meropenem-vaborbactam; PFAV, putative function-altering variant.

**Figure 4.**
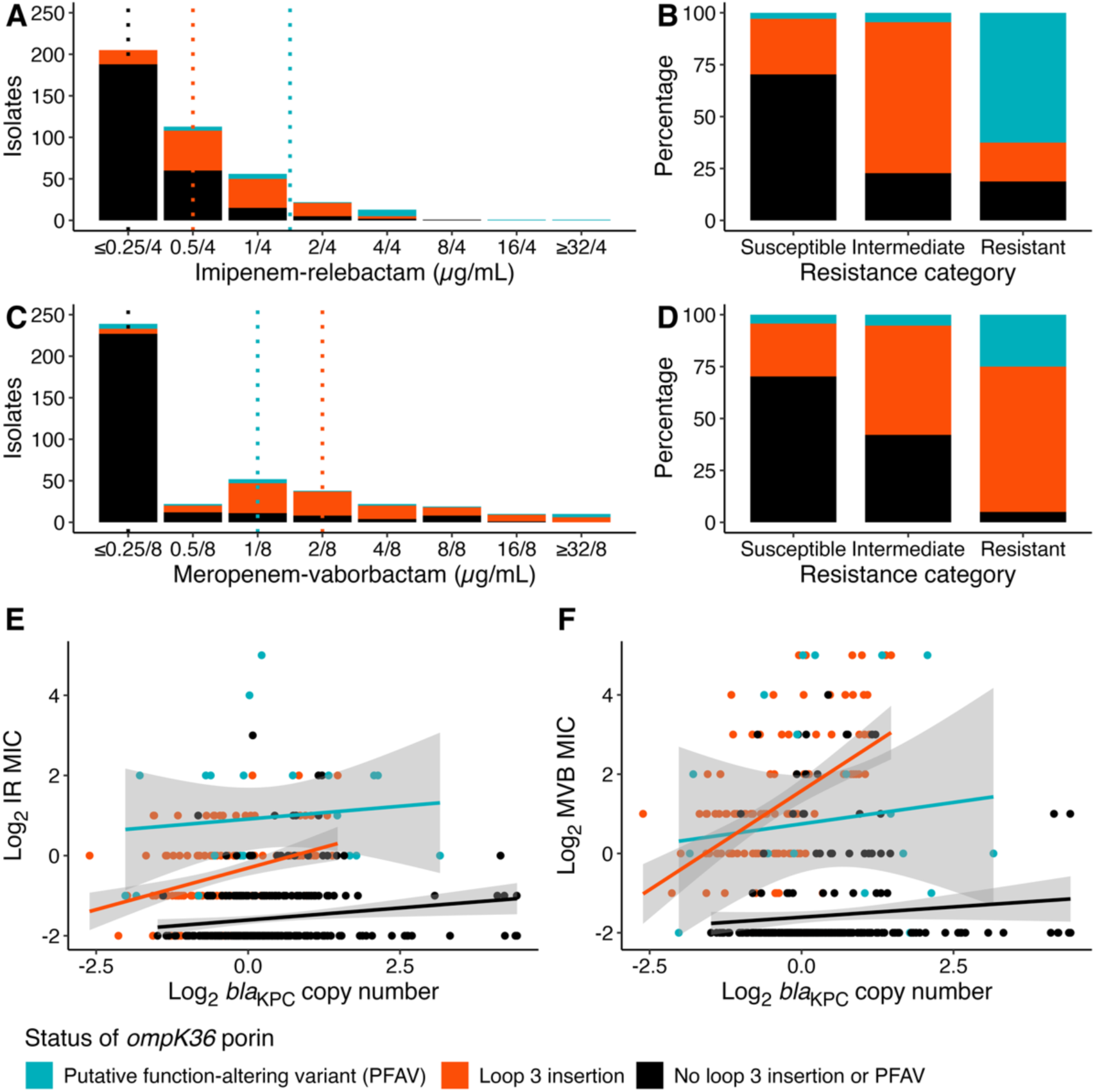
Interaction of loop 3 insertions in *ompK36* porin with *bla*_KPC_ copy number Minimum inhibitory concentration histogram stratified by the presence of mutations in the major OmpK36 porin for (**a**) imipenem-relebactam and (**c**) meropenem-vaborbactam. The 2021 Clinical and Laboratory Standards Institute breakpoints were used to categorize the susceptibility profile for (**b**) imipenem-relebactam and (**d**) meropenem-vaborbactam on stratification by the presence of mutations in the major porin gene, *ompK36*. The presence of mutations in *ompK36* porin influences the relationship between log_2_ *bla*_KPC_ copy number and log_2_ minimum inhibitory concentration for imipenem-relebactam (**e**) and meropenem-vaborbactam (**f**). **Abbreviations:** PFAV, putative function-altering variant.

The continuous GWAS identified two novel genotypes implicated in large changes to MIC. First, were putative function-altering variants in KPNIH1_RS18100, which encodes a putative efflux protein. While associated with consistent (96.3%) and large changes in MVB MIC (median log_2_ change [range], −5 [−7,7]), isolates harboring these variants had overall lower MICs than their nearest phylogenetic neighbors, leading us to not consider KPNIH1_RS18100 as an explanatory genotype. Second, were mutations in the hypothetical protein KPNIH1_RS02940, which were associated with consistent (3/5, 60%) increases in IR MIC (IR median log_2_ change [range], 3 [-1,6]). In addition to the two novel genotypes, the continuous GWAS also identified insertions in *ompK36* mapping to loop 3 of the protein, including both a composite outcome for any insertion and a specific association with GD insertions. The presence of loop 3 insertions in *ompK36*, previously reported to confer elevated carbapenem resistance^34,35^, was associated with consistent increases in MVB MIC (MVB, 93.5%) of moderate magnitude (median log_2_ fold change [range], 3 [-1,7]).

The discrete GWAS identified mutations in *ompK36* (truncation and putative function-altering variants), both of which were identified in the curated genotypic panel above. In addition, two *bla*_KPC_-containing plasmid clusters were identified as being associated with resistance, AA552 and AA018, with AA552 associated with the gain of resistance and AA018 associated with the loss of resistance. Inspection of the presence of these two plasmids on the ST258 phylogeny revealed that the gain of the *bla*_KPC_-containing AA552 plasmid was consistently associated with the loss of the AA018 plasmid, and in turn the gain of BL/BLI resistance (**Supplementary** Figure 5). Hybrid sequencing of representative isolates revealed that this pattern was due to the *bla*_KPC_ transposon, *tn4401*, hopping from the AA018 plasmid to a pre-existing AA552 plasmid, followed by loss of the AA018 plasmid backbone (**Supplementary** Figure 6). We hypothesized that this change in plasmid context mediated resistance via increased *bla*_KPC_ copy number in the AA552 plasmid, which was indeed confirmed (**Supplementary** Figure 5).

Having identified additional putative genetic mediators of resistance, we next evaluated overall explanatory power when adding these variants to the curated set. While present in three resistant isolates, KPNIH1_RS02940 mutations did not improve the explanation of resistance, as all three isolates also contained putative disruptions in *ompK36*. Loop 3 insertions in *ompK36*, while associated with increased in MIC, were lineage defining in clade I, and therefore necessary, but not sufficient to explain resistance, and therefore not considered as part of the explanatory panel. In contrast, the *bla*_KPC_-containing AA552 was both enriched in BL/BLI resistant isolates (22/58 [37.9%] vs. 4/354 [1.1%]; Fisher’s exact test p-value < 0.001) and accounted for previously unexplained resistance. Addition of this plasmid to our explanatory panel markedly improved the explanation of resistance (37.9% to 72.4%), an effect observed exclusively in clade I (17.9% to 69.2%).

The diagnostic statistics for our genotypic panel and BL/BLI resistance are reported in **Supplementary** Figure 7.

### <u>Interaction between carbapenemase copy number and loop 3 insertions in</u> *<u>ompK36</u>*

The restriction of the AA552 plasmid as a resistance-associated genotype to clade I, led us to hypothesize that that its associated increase in *bla*_KPC_ copy number exclusively modulated MIC in the clade I genetic background. Moreover, based on the observed increase in baseline MIC associated with loop 3 insertions in *ompK36*, and their restriction to clade I, we hypothesized that loop 3 insertions were necessary for AA552 to influence MIC. To test this hypothesis, we examined the association between log_2_ *bla*_KPC_ copy number and log_2_ BL/BLI MIC. Positive associations between *bla*_KPC_ copy number and MVB and IR MICs were exclusive to isolates with loop 3 insertions in *ompK36*, indicating that the restriction of porin size associated with these insertions enables the modulation of MIC through *bla*_KPC_ copy number variation. Linear regression models are reported in **Supplementary Table 2**. Together, these results suggest that the suggest that the clade I lineage defining loop 3 insertions in *ompK36* both increased baseline MIC and made accessible the evolution of resistance above the clinical breakpoint via hopping of the *bla*_KPC_-containing transposon to the higher copy number AA552 plasmid.

### Resistance-associated genotypes showed different propensities for spread

After identifying putative genetic determinants of BL/BLI resistance, we next evaluated whether the extent to which genotypes were prone to spread among patients (**Supplementary** Figure 8). Virtually all clade I resistance cluster isolates (21/24 [87.5%]) harbored the *bla*_KPC_-containing AA552 plasmid. Resistance that did not spread among patients was often mediated by convergent mutations at a small number of loci. While PBP-2 modification and putative function-altering variants in *ompK36* were observed in one small clade II clusters, these mutations were more common in resistant singletons (10/30 [33.3%]) than cluster isolates (2/28 [7.1%]) (Fisher’s exact test p-value = 0.022). Collectively, analysis of resistance-associated genotypes and BL/BLI resistance revealed that the spread of resistance was predominantly mediated by a specific *bla*_KPC_-containing plasmid, amidst a background of often dead-end mutations in genes implicated in the entry, efflux, and target of β-lactam antibiotics.

### Patient characteristics and antibiotic exposures distinguish episodes of non-carbapenemase resistance from plasmid-based resistance

Lastly, we sought to understand the role of patient characteristics and antibiotic exposures in driving the emergence and spread of resistance. Given the contribution of the *bla*_KPC_-containing AA552 plasmid to the spread of resistance, we hypothesized that distinct selective pressures influenced the acquisition of resistance via carbapenemase or non-carbapenemase mechanisms. In turn, we built exploratory logistic regression models to identify risk factors for these genotypic groupings and compared them to risk factors for harboring a resistant isolate without considering resistance genotype.

First, without accounting resistance mechanism, we found that harboring a resistant isolate was positively associated with prior exposure to cephalosporin antibiotics (adjusted odds ratio [aOR], 2.13; 95% confidence interval [CI], 1.10-4.07; p-value = 0.023), prior exposure to carbapenem antibiotics (aOR, 1.95; 95% CI, 1.03-3.67; p-value = 0.038), and increased age (adjusted odds ratio [aOR], 1.04 per 1 year unit; 95% confidence interval [95% CI], 1.02-1.07; p-value = 0.0015). A negative association with being underweight and/or malnourished was also detected (aOR, 0.42; 95% CI, 0.16-0.93; p-value = 0.047).

Next, partitioning isolates based upon their resistance genotypes and performing regression modeling revealed distinct sets of clinical associations for the acquisition of resistance through carbapenemase and non-carbapenemase mechanisms. Harboring a resistant isolate with non-carbapenemase mechanisms was positively associated with prior exposure to carbapenem antibiotics (aOR, 3.41; 95% CI, 1.52-7.77; p-value = 0.003), prior exposure to cephalosporin antibiotics (aOR, 2.63; 95% CI, 1.10-6.26; p-value = 0.028), and increased age (aOR, 1.04 per 1 year unit; 95% CI, 1.01-1.07; p-value = 0.011). Conversely, harboring a resistant isolate with the *bla*_KPC_-containing AA552 plasmid was positively associated with having chronic bronchitis and/or chronic obstructive pulmonary disease (aOR, 3.22; 95% CI, 1.11-8.92; p-value = 0.025). The unadjusted and adjusted results for all outcomes are found in **Supplementary Tables 3-5**.

Collectively, these results suggest that the contribution of antibiotic exposures to BL/BLI resistance, detected as a risk factor for harboring a resistant isolate, was mediated by the subset of patients whose isolates had dead-end mutational resistance. In contrast, restricting risk-factor analysis to transmissible resistance associated with the *bla*_KPC_-containing AA552 plasmid uncovered a potential role for patients with respiratory comorbidities in the spread of BL/BLI resistance, which was not detected when grouping all resistant isolates together.

## DISCUSSION

Amidst the growing threat of antibiotic resistance, it is critical to understand the drivers of resistance to last-line therapeutics. To this end, we leveraged bacterial whole-genome sequencing and clinical metadata from a comprehensive isolate collection to characterize the dynamics of BL/BLI resistance across a network of long-term acute care hospitals. Our analysis revealed evidence for the repeated emergence and cross-transmission of resistant strains, prior to clinical introduction of the cognate antibiotics. Placing resistance determinants in a phylogenetic context uncovered a role of genetic background in shaping the available evolutionary trajectories to resistance and identified genotypes with an elevated propensity to spread among patients. Genotypically-informed risk factor analysis implicated distinct patient characteristics and antibiotic exposures in promoting resistance through carbapenemase and non-carbapenemase mechanisms. Together, these findings have immediate implications for genomic surveillance and antibiotic stewardship efforts, alongside implications for how to identify and track the bacterial genotypes and clinical practices that promote resistance.

The discovery of pre-existing resistant strains before the clinical deployment of BL/BLI combinations is concerning. These findings align with earlier *in vitro* reports of resistance in carbapenem-resistant Enterobacterales, both before and after the availability of these therapeutics^7–13^. Further support for the generalizability of our findings comes from studies reporting similar resistance-associated genotypes when experimentally evolving carbapenem and BL/BLI resistance, even among *bla*_KPC_-containing CRKP strains^15,36,37^, and in clinical case reports of BL/BLI resistance^38–40^. Our detected association between prior carbapenem and cephalosporin exposure and the emergence of BL/BLI resistance via non-carbapenemase mechanisms indicates that mono-therapy carbapenems and cephalosporins can exert selective pressure for BL/BLI resistance among *bla*_KPC_ harboring strains^41^. Moreover, our findings also indicate that *bla*_KPC_ containing plasmids, at sufficient copy number to confer resistance in permissible genetic backgrounds, can emerge and spread in the absence of sustained antibiotic pressure. Collectively, our study suggests that early surveillance for nascent resistance, even before the clinical deployment of a drug, can reveal pre-existing resistance, as well as the bacterial genotypes, antibiotic usage patterns and patient populations with the greatest risk for promoting resistance upon clinical deployment.

Phylogenetic analysis revealed the importance of chromosome and plasmid genetic background in shaping the emergence and spread of BL/BLI resistance. In clade II, resistance predominantly emerged through dead-end chromosomal non-carbapenemase mechanisms of carbapenem resistance. In contrast, loop 3 insertions in the *ompK36* porin shaped the availability of evolutionary trajectories to resistance in clade I. Through reduced antibiotic diffusion into the cell^34,35^, this widespread insertion can synergistically interact with carbapenemses to increase resistance to carbapenems^34,42^. Consistent with this, we observe clade I isolates with loop 3 insertions had elevated baseline MICs, with transitions above the clinical breakpoint associated with transposition of the *tn4401* transposon to a plasmid context with higher copy number, a pathway previously reported to lead to resistance^43–46^. Clade I isolates with this alternative plasmid context exhibited clinical resistance that spread among patients and across numerous facilities. Collectively, these findings support the hypothesis that that chromosomal and plasmid genetic background influences the availability of resistance and potential for the clonal expansion of resistant lineages^26,28,30,34,47–51^, highlighting the importance of considering the influence of genetic background and circulating plasmids on the propensity for resistance to emerge and spread upon a therapeutic’s clinical deployment.

Genotypically-informed risk factor modeling sharpened our understanding of the patient factors that drive the emergence and spread of resistance. Evaluating risk factors for non-carbapenemase mechanisms of resistance, a common source of dead-end mutational resistance, revealed a role for antibiotic exposures in the emergence and spread of BL/BLI resistance. Specifically, prior exposure to carbapenem and cephalosporin antibiotics was positively associated with mutational pathways to BL/BLI resistance. Modeling risk factors for the acquisition of resistant strains with the *bla*_KPC_-containing AA552 plasmid revealed a positive association with the presence of chronic respiratory comorbidities (e.g., chronic bronchitis or chronic obstructive pulmonary disease). The failure to detect antibiotics as a risk factor for carrying the *bla*_KPC_-containing AA552 plasmid highlights a potential for spread among patients without sustained antibiotic exposures. Collectively, our findings suggest an interaction between bacterial genetic background, patient comorbidities, and antibiotic exposures that influence the dynamics of BL/BLI resistance.

This study has several limitations. First, the generalizability of these findings to other populations or carbapenem-resistant Enterobacterales lineages may be limited. Our analysis was restricted to *bla*_KPC_-containing ST258, the dominant sequence type in our US LTACH isolate collection^20^. Regardless, the observed chromosomal mutations and mobile genetic elements have been demonstrated to be associated with BL/BLI resistance in other strains of *K. pneumoniae*^14,15,44,52–54^, and more broadly other species of Enterobacterales, supporting the generalizability of these mechanisms^54,55^. Second, we did not evaluate whether facility-level characteristics drive the emergence or spread of resistance. Future work should evaluate the contribution of colonization burden, antibiotic stewardship, and infection prevention practices on the development and spread of resistance to BL/BLI combinations. Third, no evaluation of the necessity or sufficiency of the novel resistance-associated genotypes or putative function-altering variants in causing resistance changes was performed, although most predicted genotypes have been demonstrated to be associated with carbapenem resistance in other contexts or supported with nearest-neighbor comparison. Finally, the small sample size may have hindered our ability to differentiate risk factors for the emergence and spread of BL/BLI resistance. The application of phylogenetic contextualization in larger, densely sampled isolate collections would improve our ability to detect the patient characteristics and medical practices that distinguish these distinct situations.

In summary, we leveraged patient and genomic data to evaluate the dynamics of resistance to two promising β-lactam/β-lactamase inhibitor therapy agents before their market introduction in CRKP collected from LTACHs. Our findings demonstrate the utility of pre-introduction surveillance to illuminate the potential drivers of resistance emergence and spread within high-risk patient populations and inform future surveillance, infection prevention, and antibiotic stewardship efforts.

## METHODS

### Carbapenem-resistant *Klebsiella pneumoniae* isolate collection

Clinical isolates were collected from August 1, 2014, to July 25, 2015, at 21 United States LTACHs belonging to a national healthcare network: 12 southern California LTACHs (11 Los Angeles & 1 San Diego), six Texas LTACHs, two Florida LTACHs, and one Kentucky LTACH^20^. Bacteria from blood, respiratory, urine, and wound cultures identified as *K. pneumoniae* were tested for carbapenem resistance according to 2015 Centers for Disease Control and Prevention criteria, as described elsewhere^20^.

### Patient clinical metadata

Clinical metadata were extracted from electronic medical records, as described elsewhere^20,50^. Extracted metadata included patient demographics, medical comorbidities, and the presence of indwelling devices at specimen collection. Antibiotic exposure within the 30 days preceding isolate collection was also extracted from healthcare network records.

### Antibiotic susceptibility testing

Antibiotic susceptibility testing for newer and last-line therapeutics was performed in 2021 using a custom broth microdilution panel, as described elsewhere (SENSITITRE, Thermo Scientific)^6^. Resistance was defined as a MIC within the intermediate or resistant categories. Resistance to BL/BLI combinations was defined as having resistance to IR and/or MVB.

### Whole genome sequencing and genomic data curation

Whole-genome sequencing was performed on an Illumina HiSeq 2500 instrument, as described elsewhere^20^. Short-read genome assemblies were generated using an in-house pipeline (https://github.com/Snitkin-Lab-Umich/QCD), as described elsewhere^50^.

Reference-based variant calling was performed using an in-house pipeline (https://github.com/Snitkin-Lab-Umich/snpkit). Briefly, filtered reads were mapped to the *K. pneumoniae* KPNIH1 ST258 genome (NZ_CP008827.1) using a Burrows-Wheeler aligner v0.7.17^56^. Polymerase chain reaction duplicates were removed using PicARD v2.27.4^57^. Single-nucleotide polymorphisms (SNPs) and small insertions and deletions (INDELs) were called using SAMtools v1.2 and bcftools v1.2^58,59^.Variants were filtered using the Genome Analysis Toolkit v4.1.4.0’s VariantFiltration tool^57^. The maximum-likelihood phylogenetic tree was constructed on Gubbins-recombination filtered SNPs using a custom, in-house Snakemake pipeline (https://github.com/Snitkin-Lab-Umich/phylokit), as described elsewhere^50,60,61^.

Insertion sequences were identified using panISA v0.1.7^62^. The impact of SNPs was predicted by sequence homology using Sorting Intolerant from Tolerant (SIFT)^63,64^.

Kleborate v3.1.3 screened genome assemblies for multilocus-sequence type and antibiotic resistance determinants^65^. As over 90% of isolates belonged to the epidemic lineage ST258, our analysis focused on those isolates.

### Characterization of phylogenetic emergence and spread of bacterial phenotypes and genotypes

The phylogenetics of BL/BLI resistance and bacterial genotypes were characterized using the R package, *phyloAMR* (https://github.com/kylegontjes/phyloAMR)^50^.

*PhyloAMR’s* core function, *asr*, inferred episodes of trait gain, loss, and continuation across the phylogeny using *corHMM*’s joint ancestral state reconstruction algorithm^66^. Although the all rates different model (ARD) was the best fitting model in our original analysis of California-based isolates^50^ and this extended dataset, the equal rates (ER) model was leveraged based on its alignment with the genotypic data. In this context, the ER model better captures the observed distribution and transitions of putative resistance-associated genotypes, making it a more biologically appropriate choice for addressing the aims of this study.

The evolutionary history of a trait-containing isolate was inferred using *phyloAMR’s asr_cluster_detection()*. The phylogeny was traversed from tip to root to infer whether an isolate with the trait was a phylogenetic singleton (i.e., evidence of *de novo* evolution of the trait) or a member of a phylogenetic cluster (i.e., evidence that the trait was inherited from a circulating trait-containing lineage). Next, the synchronous transitions of BL/BLI resistance and genotypes were inferred using *phyloAMR’s synchronous_detection()*. Nodes with transitions in trait status were compared to identify instances where synchronous transitions of both traits were detected.

### Testing for different rates of BL/BLI evolution across ST258 clades

To characterize the evolution of BL/BLI resistance across ST258 clades, we leveraged *phytools*’ function, *fitmultiMK*(). This function implements a modified Markov model that permits heterogeneous rates across user-specified regions of the phylogeny^67,68^. To test the hypothesis that the evolution of BL/BLI resistance was different across clades, an ER model where rate heterogeneity was permitted on each clade was compared to an ER model with uniform rates. The fit of the single- and two-regime models was compared using the likelihood ratio test.

### Evaluating the contribution of non-carbapenemase mechanisms of carbapenem resistance to BL/BLI resistance

The fraction of BL/BLI resistance explained by non-carbapenemase mechanisms of carbapenem resistance genotypes was defined as the number of resistant isolates with the genotype divided by the total number of resistant isolates. The following genotypes were considered: *ompK35* and *ompK36* porin modification, efflux pump upregulation via mutation in regulatory genes (*ramA* and *ramR*), and modification of penicillin-binding proteins^31–33^. As the impact of resistance-associated genotypes can differ across genetic backgrounds^51^, we only considered genotypes with a median log_2_ fold change >1 using our nearest neighbor comparison algorithm, as described below.

### Nearest-neighbor comparisons to characterize a genotype’s influence on resistance

The influence of genotypes on BL/BLI resistance was characterized using *phyloAMR’s nearest_neighbor_algorithm*(). For each genotype-containing isolate, the nearest phylogenetic neighbor without the genotype was identified using ape’s *cophenetic.phylo()*. When an isolate had more than one nearest neighbor according to pairwise phylogenetic distance, the isolate with the lowest pairwise SNP distance was determined using ape’s *dist.dna*(). The MIC and BL/BLI resistance status of these pairs was compared. Summary statistics were generated to determine the magnitude and consistency of MIC changes and transitions in resistance status.

Nearest-neighbor comparisons were performed to minimize potential false-positive genotypes generated by GWAS and curate non-carbapenemase mechanisms of carbapenem resistance for inclusion in an exploratory panel of BL/BLI resistance-associated genotypes.

### Identification of *Klebsiella pneumoniae* carbapenemase-containing plasmids

MOB-suite v.3.1.9 reconstructed plasmid content from our short-read genome assemblies^69^. The *Klebsiella pneumoniae* carbapenemase, *bla*_KPC_, was identified using blastn v.2.15.0^70^ and a database of *bla*_KPC_ alleles found in the Comprehensive Antibiotic Resistance Database v.4.0.0^71^. The *bla*_KPC_-containing contigs were mapped to their inferred location (i.e., chromosome or plasmid) using MOB-recon’s contig reports.

### Genome-wide association studies to identify genomic features driving resistance

An unbiased convergence-based genome-wide association study on log_2_ MIC and BL/BLI resistance was performed using hogwash v.1.2.6^72^. Three genotype matrices were analyzed: (1) mutations in the *ompk36* porin and the presence of β-lactamase genes inferred from Kleborate, (2) *bla*_KPC_-containing plasmid clusters inferred from Mob-suite, and (3) grouped-by-gene core genome variants.

As most variants were rare (4309 variants [75.2%] had a frequency < 1%), grouped-by-gene core genome matrices were curated (**Supplementary** Figure 9). The presence of at least one variant in a genetic region (e.g., coding or non-coding region) of the reference genome was used. The following grouped-by-gene schemas were used: (1) all core genome variants identified, (2) non-synonymous coding variants (i.e., non-synonymous SNPs, coding INDELs, and coding IS), and (3) putative function-altering variants (deleterious SNPs predicted by SIFT, stop/start-modifying SNPs, frameshift INDELs, disruptive INDELs, and coding IS). A complementary analysis was performed by removing IS from our matrices. Only genotypes with a median log_2_ MIC fold change > |1| were retained for downstream analyses.

### Long-read sequencing to evaluate plasmid sharing in clade I

Representative clade I isolates underwent long-read sequencing, performed by Plasmidsaurus using Oxford Nanopore Technology, to evaluate differences among isolates with *bla*_KPC_-containing AA552 plasmids and isolates with *bla*_KPC_-containing plasmids. Hybrid assemblies were generated using an in-house assembly pipeline (https://github.com/Snitkin-Lab-Umich/Nanosake). Briefly, filtlong v0.2.1 cleaned Nanopore reads were assembled using Flye v.2.9.5^73,74^. The assembly is polished with long reads using Medaka v1.2.0, followed by polishing using clean, trimmed Illumina short-read sequences using Polypolish v.0.6.0^75^.

Whole-genome alignments of representative genomes were generated using Mummer4^76^. Dot plots graphically represented this alignment, with Blastn v2.15 used to infer the location of the *tn4401* transposon^70^.

### Copy number estimation of the *Klebsiella pneumoniae* carbapenemase gene using coverage depth

The copy number of *bla*_KPC_ was inferred using coverage statistics via an in-house Snakemake pipeline (https://github.com/kylegontjes/CoverageStats). Filtered reads were mapped to the KPNIH1 chromosome (NZ_CP008827.1) and *bla*_KPC_-containing plasmid (NZ_CP008830.1) using the Burrows-Wheeler short-read aligner v0.7.17^56^. Polymerase chain reaction duplicates were removed from sorted BAM files using SAMtools v1.19 and PicARD v.3.4.0^57,59^. The Genome Analysis Toolkit’s DepthOfCoverage v4.6.2.0 calculated coverage depth^57^. The median non-zero coverage depth was calculated for the *bla*_KPC-3_ gene (KPNIH1_RS28775) and normalized to the KPNIH1 chromosome’s median non-zero coverage depth.

Regression modeling was performed to evaluate the association between *bla_KPC_* copy number and BL/BLI resistance. First, simple linear regression models were constructed between log_2_ normalized *bla_KPC_* copy number and log_2_ MIC for MVB and IR. Next, multiple linear regression models were constructed to account for the contribution of mutations in *ompK36* to resistance. Mutations in *ompK36* was constructed as a categorical variable indicating the presence of putative-function altering variants, loop 3 insertions, or neither of these mutational classes in an isolate. Finally, interaction modeling was undertaken to evaluate the relationship between *bla_KPC_* copy number and mutations in *ompK36*.

### Regression modeling to identify patient features associated with the emergence and spread of resistance

Logistic regression was employed to identify patient features associated with resistance to BL/BLI combinations and two genotype-informed resistance classifications: resistant isolates with non-carbapenemase mechanisms of resistance and resistant isolates with *bla*_KPC_-containing AA552 plasmids. For patients with multiple isolates, their first isolate was selected unless they contributed a resistant strain, in which case the first resistant isolate was chosen. For all three outcomes, susceptible isolates served as the reference population. Patient demographics, clinical comorbidities, and antibiotic exposures were eligible for regression modeling.

The purposeful selection algorithm, with limited alterations, was employed to perform a data-driven multivariable logistic regression^77^. First, univariable logistic regression was performed on all variables.

Variables with an unadjusted p-value < 0.20 were retained as candidate variables. Next, all candidate variables were included in a logistic regression model. An iterative process was performed to remove variables if their p-value was > 0.10. After compiling this preliminary model, all initially ineligible variables were iteratively included in the pre-specified model and retained if their value was < 0.10.

### Data analysis and visualization

Analyses were performed, unless otherwise stated, using R version 4.5.0^78^. The *tidyverse* v2.0.0, *ggplot2* v3.5.2, and *ggnewscale* v0.5.1 packages were used to manipulate data, generate figures, and create scales^79^. Phylogenetic analysis and visualization were performed using *ape* v5.8-1, *phytools* v2.4-4, *ggtree* v3.16.0, *corHMM* v2.10, and *phyloAMR* v0.1.0^50,66,67,80,81^. Heatmaps were generated using *ComplexHeatmap* v2.24.0^82^. Descriptive statistics were generated using the *tableone* v0.13.2 package^83^. Multi-panel figures were generated using *cowplot* v1.1.3^84^. Code for this project can be found at https://github.com/kylegontjes/pre-intro-blbli-resistance-ms.

## Data Availability

Data produced in the present study are available upon reasonable request to the authors.

## ACKNOWLEDGMENTS

Funding sources

This work was supported by the National Institute of Allergy and Infectious Diseases (NIAID) grant 5R01AI148259. K.J.G. and E.S. were also supported by the NIAID U19AI181767. K.J.G. reports training fellowships from the National Human Genome Research Institute (T32-HG000040) and NIAID (F31-AI186288).

## Author contributions

K.J.G., E.S.S., and E.L. conceived and designed the study.

J.H., P.T., E.J.C.G., and E.L. facilitated the collection and processing of isolates.

L.L. performed culturing, DNA extraction, laboratory, and long-read sequencing work.

E.S.S. supervised research.

K.J.G., A.P., and D.B. performed computational analyses.

K.J.G. and E.S.S. wrote the manuscript with input from all co-authors.

All authors read and approved the final manuscript.

**Conflicts of interest**

Jennifer Han was affiliated with the University of Pennsylvania during conduct of this research and is currently an employee of GSK and holds shares in the company. The authors report no other conflicts of interest.

## SUPPLEMENTARY INFORMATION

**Supplementary Table 1.** Clade-specific differences in the presence of non-carbapenemase mechanisms of carbapenem resistance.

| Mechanism of interest | Overall (n=412) | Clade I (n=145) | Clade II (n=267) | p-value |
| --- | --- | --- | --- | --- |
| **Carbapenem entry** |  |  |  |  |
| *ompK35* porin truncation at 25% | 412 (100.0) | 145 (100.0) | 267 (100.0) | NA |
| *ompK36* porin | 131 (31.8) | 129 (89.0) | 2 (0.7) | <0.001 |
| Cytosine-to-thymine transition at position 25 | 124 (30.1) | 124 (85.5) | 0 (0.0) | <0.001 |
| Loop 3 insertion | 119 (28.9) | 119 (82.1) | 0 (0.0) | <0.001 |
| GD insertion | 5 (1.2) | 5 (3.4) | 0 (0.0) | 0.005 |
| Non-synonymous mutation | 22 (5.3) | 13 (9.0) | 9 (3.4) | 0.029 |
| Putative function-altering variants | 20 (4.9) | 13 (9.0) | 7 (2.6) | 0.007 |
| Truncation | 8 (1.9) | 1 (0.7) | 7 (2.6) | 0.270 |
| *ompK36* intergenic region |  |  |  |  |
| Any mutation | 31 (7.5) | 5 (3.4) | 26 (9.7) | 0.020 |
| Insertion sequences | 24 (5.8) | 1 (0.7) | 23 (8.6) | 0.001 |
| **Carbapenem efflux** |  |  |  |  |
| *acrAB-tolC* efflux pump mutant | 2 (0.5) | 0 (0.0) | 2 (0.7) | 0.543 |
| *ramR* efflux pump regulator putative function-altering variants | 27 (6.6) | 11 (7.6) | 16 (6.0) | 0.537 |
| *ramA* efflux pump activator putative function-altering variants | 10 (2.4) | 4 (2.8) | 6 (2.2) | 0.746 |
| **Carbapenem target modification** |  |  |  |  |
| Non-synonymous mutations in penicillin-binding-proteins | 9 (2.2) | 3 (2.1) | 6 (2.2) | 1.000 |
| Penicillin-binding protein-2 | 8 (1.9) | 2 (1.4) | 6 (2.2) | 0.718 |
| Penicillin-binding protein-4 | 2 (0.5) | 2 (1.4) | 0 (0.0) | 0.123 |
| **B-lactamases** |  |  |  |  |
| *bla<sub>CMY-2.v2</sub>* | 12 (2.9) | 9 (6.2) | 3 (1.1) | 0.005 |
| *bla<sub>CTX-M-14</sub>* | 8 (1.9) | 8 (5.5) | 0 (0.0) | <0.001 |
| *bla<sub>CTX-M-15</sub>* | 11 (2.7) | 10 (6.9) | 1 (0.4) | <0.001 |
| *bla<sub>KPC-2</sub>* | 136 (33.0) | 131 (90.3) | 5 (1.9) | <0.001 |
| *bla<sub>KPC-3</sub>* | 269 (65.3) | 9 (6.2) | 260 (97.4) | <0.001 |
| *bla<sub>KPC-5</sub>* | 3 (0.7) | 3 (2.1) | 0 (0.0) | 0.043 |
| *bla<sub>LAP-2</sub>* | 1 (0.2) | 0 (0.0) | 1 (0.4) | 1.000 |
| *bla<sub>OXA-1</sub>* | 10 (2.4) | 10 (6.9) | 0 (0.0) | <0.001 |
| *bla<sub>OXA-9.v1</sub>* | 154 (37.4) | 1 (0.7) | 153 (57.3) | <0.001 |
| *bla<sub>SHV-11</sub>* | 313 (76.0) | 135 (93.1) | 178 (66.7) | <0.001 |
| *bla<sub>SHV-12</sub>* | 30 (7.3) | 2 (1.4) | 28 (10.5) | 0.001 |
| *bla<sub>TEM-104?</sub>* | 6 (1.5) | 6 (4.1) | 0 (0.0) | 0.002 |
| *bla<sub>TEM-104\*?</sub>* | 7 (1.7) | 6 (4.1) | 1 (0.4) | 0.009 |
| *bla<sub>TEM-150\*?</sub>* | 6 (1.5) | 0 (0.0) | 6 (2.2) | 0.095 |
| *bla<sub>TEM-1D.v1</sub>* | 177 (43.0) | 51 (35.2) | 126 (47.2) | 0.024 |
| ***tn4401* bla<sub>KPC</sub> transposon isoform** |  |  |  |  |
| *tn4401a* | 143 (35.0) | 138 (95.8) | 5 (1.9) | <0.001 |
| *tn4401b* | 39 (9.5) | 5 (3.5) | 34 (12.8) | 0.004 |
| *tn4401d* | 137 (33.5) | 1 (0.7) | 136 (51.3) | <0.001 |
| *tn4401 del 6920-7126* | 50 (12.2) | 0 (0.0) | 50 (18.9) | <0.001 |
| *tn4401 del 1-554 7008-7075* | 39 (9.5) | 0 (0.0) | 39 (14.7) | <0.001 |
| *tn4401 del 1-3391 6920-7126* | 1 (0.2) | 0 (0.0) | 1 (0.4) | 1.000 |

Statistical significance was tested using the Chi-squared test for most variables. Fisher’s exact test was performed for the following variables due to small event size: TD insertions in *ompk36* porin, *acrAB-tolC* efflux pump variant, *ramA*, *ramR*, penicillin-binding protein-2, penicillin-binding protein-4, *bla*<sub>CTX-M14</sub>, *bla*<sub>CTX-M-15</sub>, *bla*<sub>KPC-5</sub>, *bla*<sub>LAP-2</sub>, *bla*<sub>OXA-1</sub>, *bla*<sub>TEM-1</sub>, and *tn4401 del 1-3391 5920-7126*. For β-lactamase genes, ? indicates coverage < 100%, whereas \*? indicates an incomplete match and coverage <100%.

**Supplementary Table 2.**
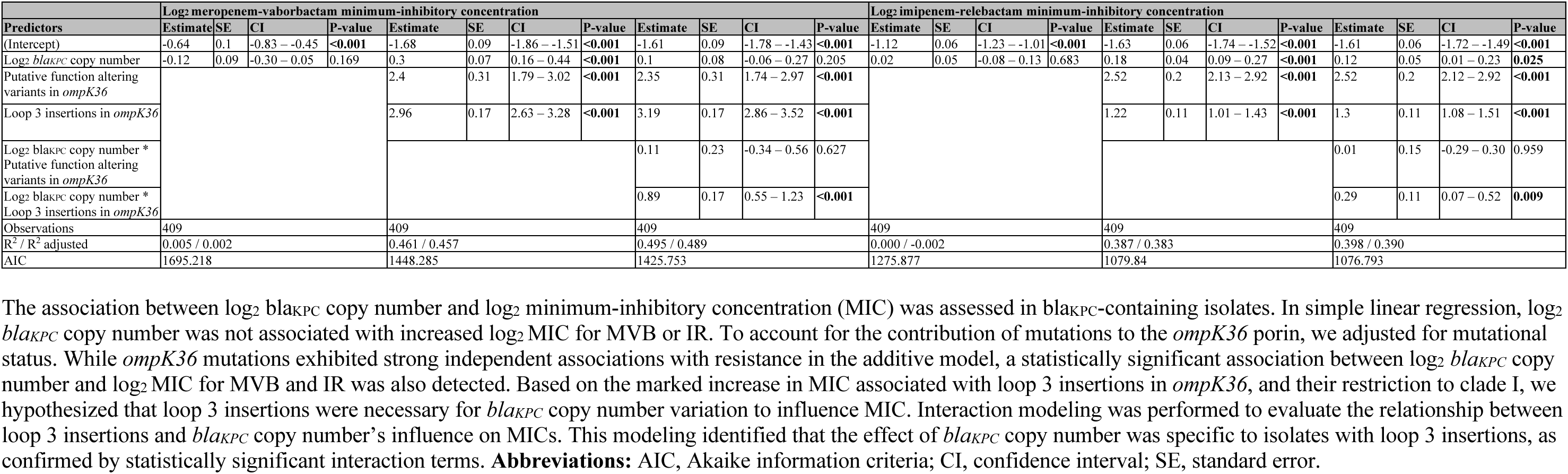
Regression modeling to evaluate interaction between loop 3 insertions in *ompK36* porin with *bla*_KPC_ copy number.

| Predictors | Log<sub>2</sub> meropenem-vaborbactam minimum-inhibitory concentration |  |  |  |  |  |  |  |  |  |  |  | Log<sub>2</sub> imipenem-relebactam minimum-inhibitory concentration |  |  |  |  |  |  |  |  |  |  |  |
| --- | --- | --- | --- | --- | --- | --- | --- | --- | --- | --- | --- | --- | --- | --- | --- | --- | --- | --- | --- | --- | --- | --- | --- | --- |
|  | Estimate | SE | CI | P-value | Estimate | SE | CI | P-value | Estimate | SE | CI | P-value | Estimate | SE | CI | P-value | Estimate | SE | CI | P-value | Estimate | SE | CI | P-value |
| (Intercept) | -0.64 | 0.1 | -0.83 – -0.45 | <0.001 | -1.68 | 0.09 | -1.86 – -1.51 | <0.001 | -1.61 | 0.09 | -1.78 – -1.43 | <0.001 | -1.12 | 0.06 | -1.23 – -1.01 | <0.001 | -1.63 | 0.06 | -1.74 – -1.52 | <0.001 | -1.61 | 0.06 | -1.72 – -1.49 | <0.001 |
| Log<sub>2</sub> *blaKPC* copy number | -0.12 | 0.09 | -0.30 – 0.05 | 0.169 | 0.3 | 0.07 | 0.16 – 0.44 | <0.001 | 0.1 | 0.08 | -0.06 – 0.27 | 0.205 | 0.02 | 0.05 | -0.08 – 0.13 | 0.683 | 0.18 | 0.04 | 0.09 – 0.27 | <0.001 | 0.12 | 0.05 | 0.01 – 0.23 | 0.025 |
| Putative function altering variants in *ompK36* |  |  |  |  | 2.4 | 0.31 | 1.79 – 3.02 | <0.001 | 2.35 | 0.31 | 1.74 – 2.97 | <0.001 |  |  |  |  | 2.52 | 0.2 | 2.13 – 2.92 | <0.001 | 2.52 | 0.2 | 2.12 – 2.92 | <0.001 |
| Loop 3 insertions in *ompK36* |  |  |  |  | 2.96 | 0.17 | 2.63 – 3.28 | <0.001 | 3.19 | 0.17 | 2.86 – 3.52 | <0.001 |  |  |  |  | 1.22 | 0.11 | 1.01 – 1.43 | <0.001 | 1.3 | 0.11 | 1.08 – 1.51 | <0.001 |
| Log<sub>2</sub> *blaKPC* copy number * Putative function altering variants in *ompK36* |  |  |  |  |  |  |  |  | 0.11 | 0.23 | -0.34 – 0.56 | 0.627 |  |  |  |  |  |  |  |  | 0.01 | 0.15 | -0.29 – 0.30 | 0.959 |
| Log<sub>2</sub> *blaKPC* copy number * Loop 3 insertions in *ompK36* |  |  |  |  |  |  |  |  | 0.89 | 0.17 | 0.55 – 1.23 | <0.001 |  |  |  |  |  |  |  |  | 0.29 | 0.11 | 0.07 – 0.52 | 0.009 |
| Observations | 409 |  |  |  | 409 |  |  |  | 409 |  |  |  | 409 |  |  |  | 409 |  |  |  | 409 |  |  |  |
| R<sup>2</sup> / R<sup>2</sup> adjusted | 0.005 / 0.002 |  |  |  | 0.461 / 0.457 |  |  |  | 0.495 / 0.489 |  |  |  | 0.000 / -0.002 |  |  |  | 0.387 / 0.383 |  |  |  | 0.398 / 0.390 |  |  |  |
| AIC | 1695.218 |  |  |  | 1448.285 |  |  |  | 1425.753 |  |  |  | 1275.877 |  |  |  | 1079.84 |  |  |  | 1076.793 |  |  |  |

The association between log<sub>2</sub> *blaKPC* copy number and log<sub>2</sub> minimum-inhibitory concentration (MIC) was assessed in *blaKPC*-containing isolates. In simple linear regression, log<sub>2</sub> *blaKPC* copy number was not associated with increased log<sub>2</sub> MIC for MVB or IR. To account for the contribution of mutations to the *ompK36* porin, we adjusted for mutational status. While *ompK36* mutations exhibited strong independent associations with resistance in the additive model, a statistically significant association between log<sub>2</sub> *blaKPC* copy number and log<sub>2</sub> MIC for MVB and IR was also detected. Based on the marked increase in MIC associated with loop 3 insertions in *ompK36*, and their restriction to clade I, we hypothesized that loop 3 insertions were necessary for *blaKPC* copy number variation to influence MIC. Interaction modeling was performed to evaluate the relationship between loop 3 insertions and *blaKPC* copy number's influence on MICs. This modeling identified that the effect of *blaKPC* copy number was specific to isolates with loop 3 insertions, as confirmed by statistically significant interaction terms. **Abbreviations:** AIC, Akaike information criteria; CI, confidence interval; SE, standard error.

**Supplementary Table 3.** Unadjusted and adjusted analysis of risk factors for resistance to beta-lactam/beta-lactamase combinations.

| Variable | Overall (n=336) | Resistant (n=52) | Susceptible (n=284) | OR (95% CI) | OR p-value | aOR (95% CI) | aOR p-value |
| --- | --- | --- | --- | --- | --- | --- | --- |
| Age. 1 year unit | 71.06 (14.66) | 77.00 (11.72) | 69.98 (14.90) | 1.04 (1.02-1.07) | 0.0017 | 1.04 (1.02-1.07) | 0.0015 |
| Female sex | 40.99 (110.33) | 41.81 (66.94) | 40.85 (116.63) | 1.00 (1.00-1.00) | 0.9538 |  |  |
| Length of stay before culture, 1-day unit | 160 (47.6) | 28 (53.8) | 132 (46.5) | 1.34 (0.74-2.45) | 0.3292 |  |  |
| Tracheostomy tube | 227 (67.6) | 33 (63.5) | 194 (68.3) | 0.81 (0.44-1.52) | 0.4929 |  |  |
| Central venous catheter | 187 (55.7) | 25 (48.1) | 162 (57.0) | 0.70 (0.38-1.26) | 0.2331 |  |  |
| Indwelling urinary catheter | 183 (54.5) | 29 (55.8) | 154 (54.2) | 1.06 (0.59-1.94) | 0.8372 |  |  |
| Gastrostomy tube | 121 (36.0) | 16 (30.8) | 105 (37.0) | 0.76 (0.39-1.41) | 0.3927 |  |  |
| Acute kidney injury | 154 (45.8) | 23 (44.2) | 131 (46.1) | 0.93 (0.51-1.68) | 0.8009 |  |  |
| Chronic kidney disease | 110 (32.7) | 18 (34.6) | 92 (32.4) | 1.10 (0.58-2.04) | 0.7537 |  |  |
| Ventilator-dependent respiratory failure | 99 (29.5) | 14 (26.9) | 85 (29.9) | 0.86 (0.43-1.64) | 0.6622 |  |  |
| Underweight or malnutrition | 84 (25.0) | 7 (13.5) | 77 (27.1) | 0.42 (0.17-0.91) | 0.0415 | 0.42 (0.16-0.93) | 0.0466 |
| Congestive heart failure | 64 (19.0) | 9 (17.3) | 55 (19.4) | 0.87 (0.38-1.82) | 0.7283 |  |  |
| Decubitus ulcer stage 4+ | 67 (19.9) | 7 (13.5) | 60 (21.1) | 0.58 (0.23-1.28) | 0.2079 |  |  |
| Chronic bronchitis or chronic obstructive pulmonary disorder | 60 (17.9) | 13 (25.0) | 47 (16.5) | 1.68 (0.81-3.32) | 0.1467 |  |  |
| Brain injury | 63 (18.8) | 11 (21.2) | 52 (18.3) | 1.20 (0.55-2.42) | 0.6294 |  |  |
| Malignancy | 42 (12.5) | 6 (11.5) | 36 (12.7) | 0.90 (0.33-2.12) | 0.8197 |  |  |
| Obesity | 19 (5.7) | 2 (3.8) | 17 (6.0) | 0.63 (0.10-2.28) | 0.5425 |  |  |
| Carbapenem exposure | 106 (31.5) | 23 (44.2) | 83 (29.2) | 1.92 (1.04-3.51) | 0.0342 | 1.95 (1.03-3.67) | 0.0383 |
| Amikacin exposure | 41 (12.2) | 8 (15.4) | 33 (11.6) | 1.38 (0.56-3.06) | 0.4473 |  |  |
| Gentamicin exposure | 12 (3.6) | 2 (3.8) | 10 (3.5) | 1.10 (0.17-4.32) | 0.9076 |  |  |
| Tobramycin exposure | 21 (6.2) | 5 (9.6) | 16 (5.6) | 1.78 (0.56-4.80) | 0.2813 |  |  |
| Aztreonam exposure | 10 (3.0) | 0 (0.0) | 10 (3.5) | 0.00 (NA-Infinity) | 0.9843 |  |  |
| Sulfamethoxazole-trimethoprim exposure | 11 (3.3) | 2 (3.8) | 9 (3.2) | 1.22 (0.18-4.92) | 0.8011 |  |  |
| Cephalosporin exposure | 83 (24.7) | 20 (38.5) | 63 (22.2) | 2.19 (1.16-4.07) | 0.0138 | 2.13 (1.10-4.07) | 0.0232 |
| Fluoroquinolone exposure | 59 (17.6) | 7 (13.5) | 52 (18.3) | 0.69 (0.27-1.54) | 0.4003 |  |  |
| Daptomycin exposure | 14 (4.2) | 5 (9.6) | 9 (3.2) | 3.25 (0.96-9.84) | 0.042 |  |  |
| Metronidazole exposure | 62 (18.5) | 10 (19.2) | 52 (18.3) | 1.06 (0.48-2.18) | 0.875 |  |  |
| Linezolid exposure | 48 (14.3) | 6 (11.5) | 42 (14.8) | 0.75 (0.27-1.75) | 0.5392 |  |  |
| Polymyxin and/or colistin exposure | 38 (11.3) | 6 (11.5) | 32 (11.3) | 1.03 (0.37-2.44) | 0.9548 |  |  |
| Tigecycline exposure | 47 (14.0) | 11 (21.2) | 36 (12.7) | 1.85 (0.84-3.83) | 0.1093 |  |  |
| Vancomycin exposure | 143 (42.6) | 21 (40.4) | 122 (43.0) | 0.90 (0.49-1.63) | 0.7302 |  |  |
| Piperacillin/Tazobactam exposure | 45 (13.4) | 5 (9.6) | 40 (14.1) | 0.65 (0.22-1.59) | 0.3875 |  |  |

**Abbreviations:** aOR, adjusted odds ratio; CI, confidence interval; OR, odds ratio

**Supplementary Table 4.** Unadjusted and adjusted analysis of risk factors for non-carbapenemase mechanisms of resistance to beta-lactam/beta-lactamase combinations.

| Variable | Overall (n=317) | Resistant (n=34) | Susceptible (n=284) | OR (95% CI) | OR p-value | aOR (95% CI) | aOR p-value |
| --- | --- | --- | --- | --- | --- | --- | --- |
| Age. 1 year unit | 70.75 (14.84) | 77.20 (12.84) | 69.98 (14.90) | 1.04 (1.01-1.07) | 0.0079 | 1.04 (1.01-1.07) | 0.0112 |
| Female sex | 40.46 (110.78) | 37.26 (35.17) | 40.85 (116.63) | 1.00 (0.99-1.00) | 0.8591 |  |  |
| Length of stay before culture, 1-day unit | 149 (46.9) | 17 (50.0) | 132 (46.5) | 1.15 (0.56-2.36) | 0.6976 |  |  |
| Tracheostomy tube | 217 (68.2) | 23 (67.6) | 194 (68.3) | 0.97 (0.46-2.15) | 0.9375 |  |  |
| Central venous catheter | 178 (56.0) | 16 (47.1) | 162 (57.0) | 0.67 (0.32-1.37) | 0.2701 |  |  |
| Indwelling urinary catheter | 170 (53.5) | 16 (47.1) | 154 (54.2) | 0.75 (0.36-1.53) | 0.4296 |  |  |
| Gastrostomy tube | 114 (35.8) | 9 (26.5) | 105 (37.0) | 0.61 (0.26-1.32) | 0.2311 |  |  |
| Acute kidney injury | 143 (45.0) | 12 (35.3) | 131 (46.1) | 0.64 (0.30-1.32) | 0.2331 |  |  |
| Chronic kidney disease | 103 (32.4) | 11 (32.4) | 92 (32.4) | 1.00 (0.45-2.09) | 0.9961 |  |  |
| Ventilator-dependent respiratory failure | 93 (29.2) | 8 (23.5) | 85 (29.9) | 0.72 (0.29-1.59) | 0.4398 |  |  |
| Underweight or malnutrition | 81 (25.5) | 4 (11.8) | 77 (27.1) | 0.36 (0.10-0.95) | 0.0615 | 0.40 (0.11-1.09) | 0.1012 |
| Congestive heart failure | 59 (18.6) | 4 (11.8) | 55 (19.4) | 0.56 (0.16-1.48) | 0.2873 |  |  |
| Decubitus ulcer stage 4+ | 64 (20.1) | 4 (11.8) | 60 (21.1) | 0.50 (0.14-1.32) | 0.2061 |  |  |
| Chronic bronchitis or chronic obstructive pulmonary disorder | 53 (16.7) | 6 (17.6) | 47 (16.5) | 1.08 (0.39-2.60) | 0.8711 |  |  |
| Brain injury | 58 (18.2) | 6 (17.6) | 52 (18.3) | 0.96 (0.34-2.28) | 0.9247 |  |  |
| Malignancy | 39 (12.3) | 3 (8.8) | 36 (12.7) | 0.67 (0.15-1.99) | 0.5201 |  |  |
| Obesity | 18 (5.7) | 1 (2.9) | 17 (6.0) | 0.48 (0.03-2.44) | 0.4776 |  |  |
| Carbapenem exposure | 100 (31.4) | 17 (50.0) | 83 (29.2) | 2.42 (1.17-5.00) | 0.0159 | 3.41 (1.52-7.77) | 0.0031 |
| Amikacin exposure | 38 (11.9) | 5 (14.7) | 33 (11.6) | 1.31 (0.42-3.37) | 0.601 |  |  |
| Gentamicin exposure | 12 (3.8) | 2 (5.9) | 10 (3.5) | 1.71 (0.26-6.87) | 0.4996 |  |  |
| Tobramycin exposure | 20 (6.3) | 4 (11.8) | 16 (5.6) | 2.23 (0.61-6.57) | 0.1741 |  |  |
| Aztreonam exposure | 10 (3.1) | 0 (0.0) | 10 (3.5) | 0.00 (NA-Infinity) | 0.9901 |  |  |
| Sulfamethoxazole-trimethoprim exposure | 10 (3.1) | 1 (2.9) | 9 (3.2) | 0.93 (0.05-5.16) | 0.9427 |  |  |
| Cephalosporin exposure | 76 (23.9) | 13 (38.2) | 63 (22.2) | 2.17 (1.01-4.54) | 0.0417 | 2.63 (1.10-6.26) | 0.0281 |
| Fluoroquinolone exposure | 57 (17.9) | 5 (14.7) | 52 (18.3) | 0.77 (0.25-1.93) | 0.6055 |  |  |
| Daptomycin exposure | 12 (3.8) | 3 (8.8) | 9 (3.2) | 2.96 (0.63-10.52) | 0.1177 |  |  |
| Metronidazole exposure | 57 (17.9) | 5 (14.7) | 52 (18.3) | 0.77 (0.25-1.93) | 0.6055 |  |  |
| Linezolid exposure | 44 (13.8) | 2 (5.9) | 42 (14.8) | 0.36 (0.06-1.25) | 0.172 | 0.26 (0.04-0.99) | 0.0878 |
| Polymyxin and/or colistin exposure | 37 (11.6) | 5 (14.7) | 32 (11.3) | 1.36 (0.44-3.50) | 0.5559 |  |  |
| Tigecycline exposure | 45 (14.2) | 9 (26.5) | 36 (12.7) | 2.48 (1.03-5.58) | 0.0337 |  |  |
| Vancomycin exposure | 135 (42.5) | 13 (38.2) | 122 (43.0) | 0.82 (0.39-1.69) | 0.5989 | 0.46 (0.18-1.11) | 0.0914 |
| Piperacillin/Tazobactam exposure | 43 (13.5) | 3 (8.8) | 40 (14.1) | 0.59 (0.14-1.76) | 0.4015 |  |  |

**Abbreviations:** aOR, adjusted odds ratio; CI, confidence interval; OR, odds ratio.

**Supplementary Table 5.** Unadjusted and adjusted analysis of risk factors for plasmid-mediated mechanisms of resistance to beta-lactam/beta-lactamase combinations.

| Variable | Overall (n=302) | Resistant (n=18) | Susceptible (n=284) | OR (95% CI) | OR p-value | aOR (95% CI) | aOR p-value |
| --- | --- | --- | --- | --- | --- | --- | --- |
| Age. 1 year unit | 70.37 (14.71) | 76.63 (9.58) | 69.98 (14.90) | 1.04 (1.00-1.08) | 0.0633 | 1.04 (1.00-1.08) | 0.0771 |
| Female sex | 41.41 (115.80) | 50.39 (104.52) | 40.85 (116.63) | 1.00 (0.99-1.00) | 0.7389 |  |  |
| Length of stay before culture, 1-day unit | 143 (47.4) | 11 (61.1) | 132 (46.5) | 1.81 (0.69-5.04) | 0.2336 |  |  |
| Tracheostomy tube | 204 (67.5) | 10 (55.6) | 194 (68.3) | 0.58 (0.22-1.57) | 0.2673 |  |  |
| Central venous catheter | 171 (56.6) | 9 (50.0) | 162 (57.0) | 0.75 (0.29-1.98) | 0.5599 |  |  |
| Indwelling urinary catheter | 167 (55.3) | 13 (72.2) | 154 (54.2) | 2.19 (0.80-6.99) | 0.1451 |  |  |
| Gastrostomy tube | 112 (37.1) | 7 (38.9) | 105 (37.0) | 1.08 (0.39-2.84) | 0.8703 |  |  |
| Acute kidney injury | 142 (47.0) | 11 (61.1) | 131 (46.1) | 1.84 (0.70-5.11) | 0.2226 |  |  |
| Chronic kidney disease | 99 (32.8) | 7 (38.9) | 92 (32.4) | 1.33 (0.48-3.48) | 0.5703 |  |  |
| Ventilator-dependent respiratory failure | 91 (30.1) | 6 (33.3) | 85 (29.9) | 1.17 (0.40-3.12) | 0.7604 |  |  |
| Underweight or malnutrition | 80 (26.5) | 3 (16.7) | 77 (27.1) | 0.54 (0.12-1.68) | 0.3371 |  |  |
| Congestive heart failure | 60 (19.9) | 5 (27.8) | 55 (19.4) | 1.60 (0.50-4.44) | 0.3895 |  |  |
| Decubitus ulcer stage 4+ | 63 (20.9) | 3 (16.7) | 60 (21.1) | 0.75 (0.17-2.35) | 0.6526 |  |  |
| Chronic bronchitis or chronic obstructive pulmonary disorder | 54 (17.9) | 7 (38.9) | 47 (16.5) | 3.21 (1.13-8.58) | 0.022 | 3.22 (1.11-8.92) | 0.0254 |
| Brain injury | 57 (18.9) | 5 (27.8) | 52 (18.3) | 1.72 (0.53-4.77) | 0.3246 |  |  |
| Malignancy | 39 (12.9) | 3 (16.7) | 36 (12.7) | 1.38 (0.31-4.43) | 0.6258 |  |  |
| Obesity | 18 (6.0) | 1 (5.6) | 17 (6.0) | 0.92 (0.05-4.94) | 0.9404 |  |  |
| Carbapenem exposure | 89 (29.5) | 6 (33.3) | 83 (29.2) | 1.21 (0.41-3.23) | 0.7112 |  |  |
| Amikacin exposure | 36 (11.9) | 3 (16.7) | 33 (11.6) | 1.52 (0.34-4.92) | 0.5244 |  |  |
| Gentamicin exposure | 10 (3.3) | 0 (0.0) | 10 (3.5) | 0.00 (NA-Infinity) | 0.9905 |  |  |
| Tobramycin exposure | 17 (5.6) | 1 (5.6) | 16 (5.6) | 0.99 (0.05-5.30) | 0.9889 |  |  |
| Aztreonam exposure | 10 (3.3) | 0 (0.0) | 10 (3.5) | 0.00 (NA-Infinity) | 0.9905 |  |  |
| Sulfamethoxazole-trimethoprim exposure | 10 (3.3) | 1 (5.6) | 9 (3.2) | 1.80 (0.09-10.42) | 0.5883 |  |  |
| Cephalosporin exposure | 70 (23.2) | 7 (38.9) | 63 (22.2) | 2.23 (0.79-5.91) | 0.1112 | 2.66 (0.92-7.36) | 0.0606 |
| Fluoroquinolone exposure | 54 (17.9) | 2 (11.1) | 52 (18.3) | 0.56 (0.09-2.04) | 0.4456 |  |  |
| Daptomycin exposure | 11 (3.6) | 2 (11.1) | 9 (3.2) | 3.82 (0.55-16.44) | 0.1034 |  |  |
| Metronidazole exposure | 57 (18.9) | 5 (27.8) | 52 (18.3) | 1.72 (0.53-4.77) | 0.3246 |  |  |
| Linezolid exposure | 46 (15.2) | 4 (22.2) | 42 (14.8) | 1.65 (0.45-4.85) | 0.399 |  |  |
| Polymyxin and/or colistin exposure | 33 (10.9) | 1 (5.6) | 32 (11.3) | 0.46 (0.03-2.38) | 0.4619 |  |  |
| Tigecycline exposure | 38 (12.6) | 2 (11.1) | 36 (12.7) | 0.86 (0.13-3.20) | 0.8462 |  |  |
| Vancomycin exposure | 130 (43.0) | 8 (44.4) | 122 (43.0) | 1.06 (0.39-2.77) | 0.9017 |  |  |
| Piperacillin/Tazobactam exposure | 42 (13.9) | 2 (11.1) | 40 (14.1) | 0.76 (0.12-2.82) | 0.7244 |  |  |

**Abbreviations:** aOR, adjusted odds ratio; CI, confidence interval; OR, odds ratio.

**Supplementary Figure 1.**
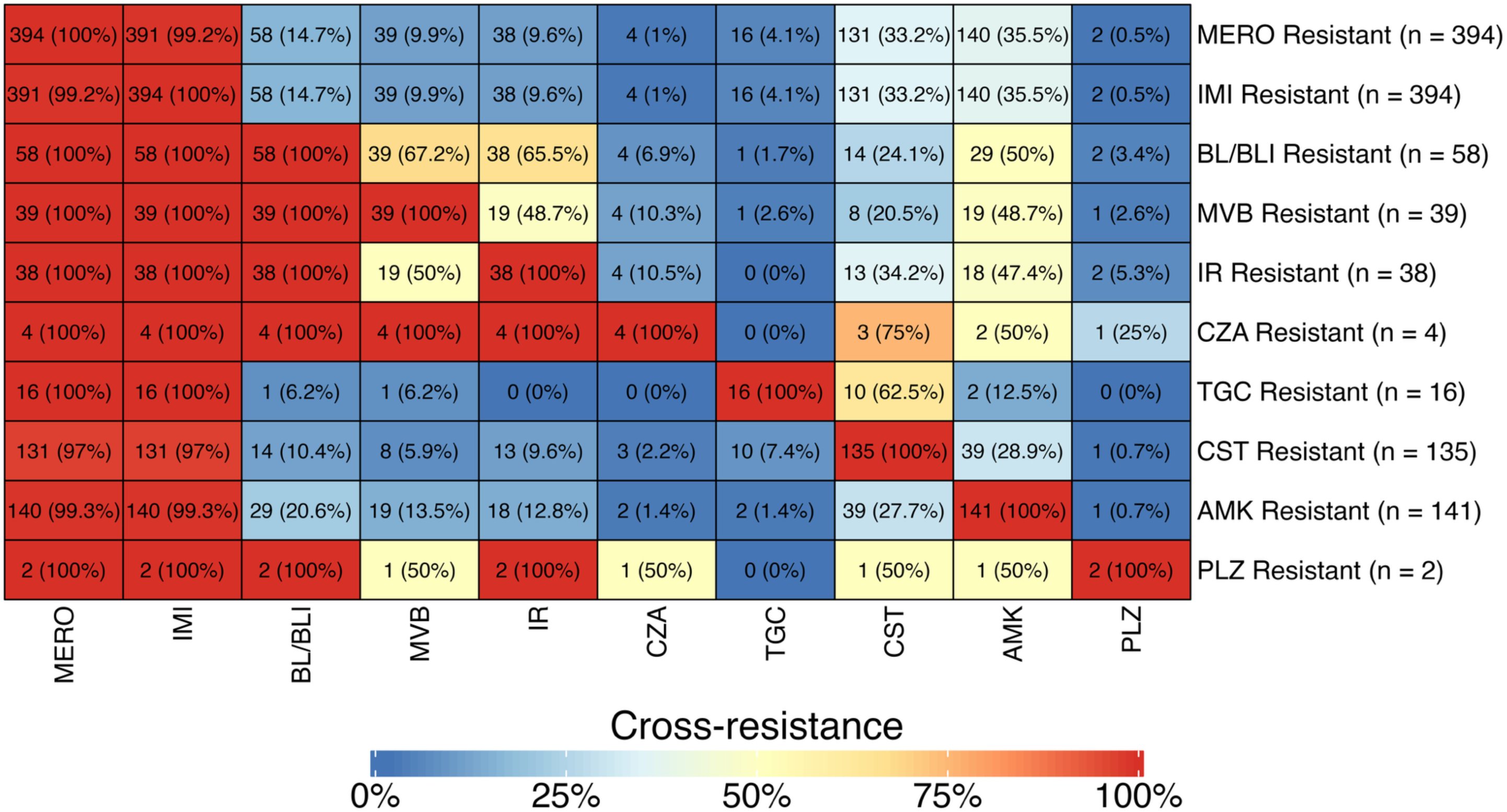
Cross-resistance profile of carbapenem-resistant *Klebsiella pneumoniae* ST258 isolates The cross-resistance profile of 412 carbapenem-resistant *Klebsiella pneumoniae* sequence type 258 strains. This heatmap shows the number of resistant isolates (rows) that also exhibit resistance to other antibiotics (columns). Cross-resistance was evaluated for the following antibiotics: amikacin, colistin, ceftazidime-avibactam, imipenem, imipenem-relebactam, meropenem, meropenem-vaborbactam, plazomicin, and tigecycline. Resistance was defined as a minimum inhibitory concentration within intermediate or resistant categories. Resistance to β-lactam/β-lactamase inhibitor combinations was defined as having resistance to imipenem-relebactam and/or meropenem-vaborbactam. **Abbreviations:** AMK, amikacin; BL/BLI, β-lactam/β-lactamase inhibitor; CST, colistin; CZA, ceftazidime-avibactam; IMI, imipenem; IR, imipenem-relebactam; MERO, meropenem, MVB, meropenem-vaborbactam; PLZ, plazomicin; TGC, tigecycline.

**Supplementary Figure 2.**
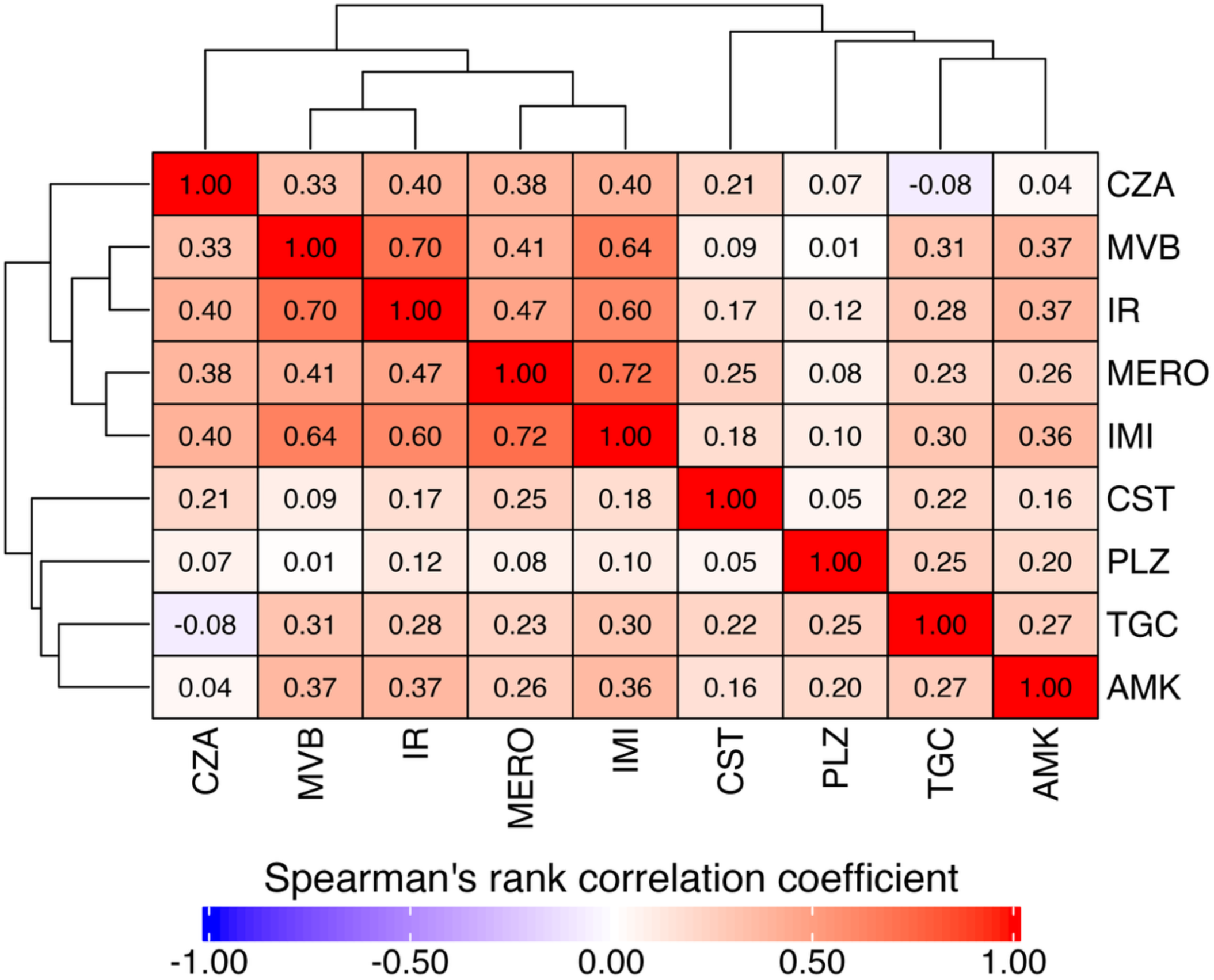
Spearman’s rank correlation coefficients between the minimum inhibitory concentration of last-line and novel agents Spearman’s rank correlation coefficient between minimum-inhibitory concentrations of last-line and novel agents for 412 carbapenem-resistant *Klebsiella pneumoniae* sequence type 258 strains. The following antibiotics were evaluated: amikacin, colistin, ceftazidime-avibactam, imipenem, imipenem-relebactam, meropenem, meropenem-vaborbactam, plazomicin, and tigecycline. **Abbreviations:** AMK, amikacin; CST, colistin; CZA, ceftazidime-avibactam; IMI, imipenem; IR, imipenem-relebactam; MERO, meropenem, MVB, meropenem-vaborbactam; PLZ, plazomicin; TGC, tigecycline.

**Supplementary Figure 3.**
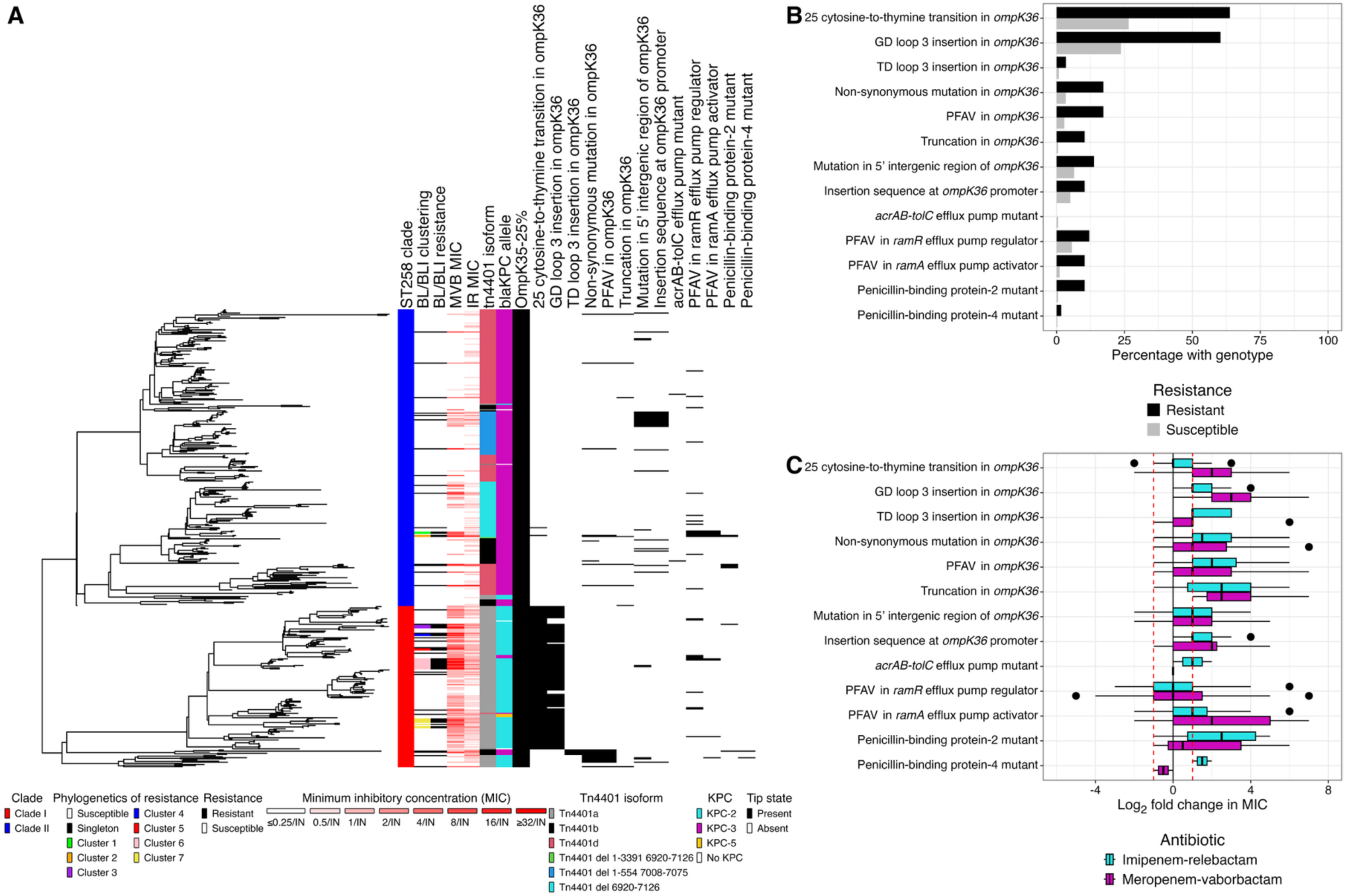
Clade-specific differences in non-carbapenemase mechanisms of carbapenem resistance (**a**) Mechanisms of carbapenem resistance were overlaid across the sequence type 258 phylogeny. (**b**) The frequency of genotypes in β-lactam/β-lactamase inhibitor combination susceptible and resistant isolates. (**c**) The log_2_ fold change in imipenem-relebactam and meropenem-vaborbactam minimum inhibitory concentration for isolates with the genotype relative to their nearest phylogenetic neighbor without the genotype. **Abbreviations:** BL/BLI, β-lactam/β-lactamase inhibitor; IR, imipenem-relebactam; MVB, meropenem-vaborbactam; PFAV, putative function-altering variant.

**Supplementary Figure 4.**
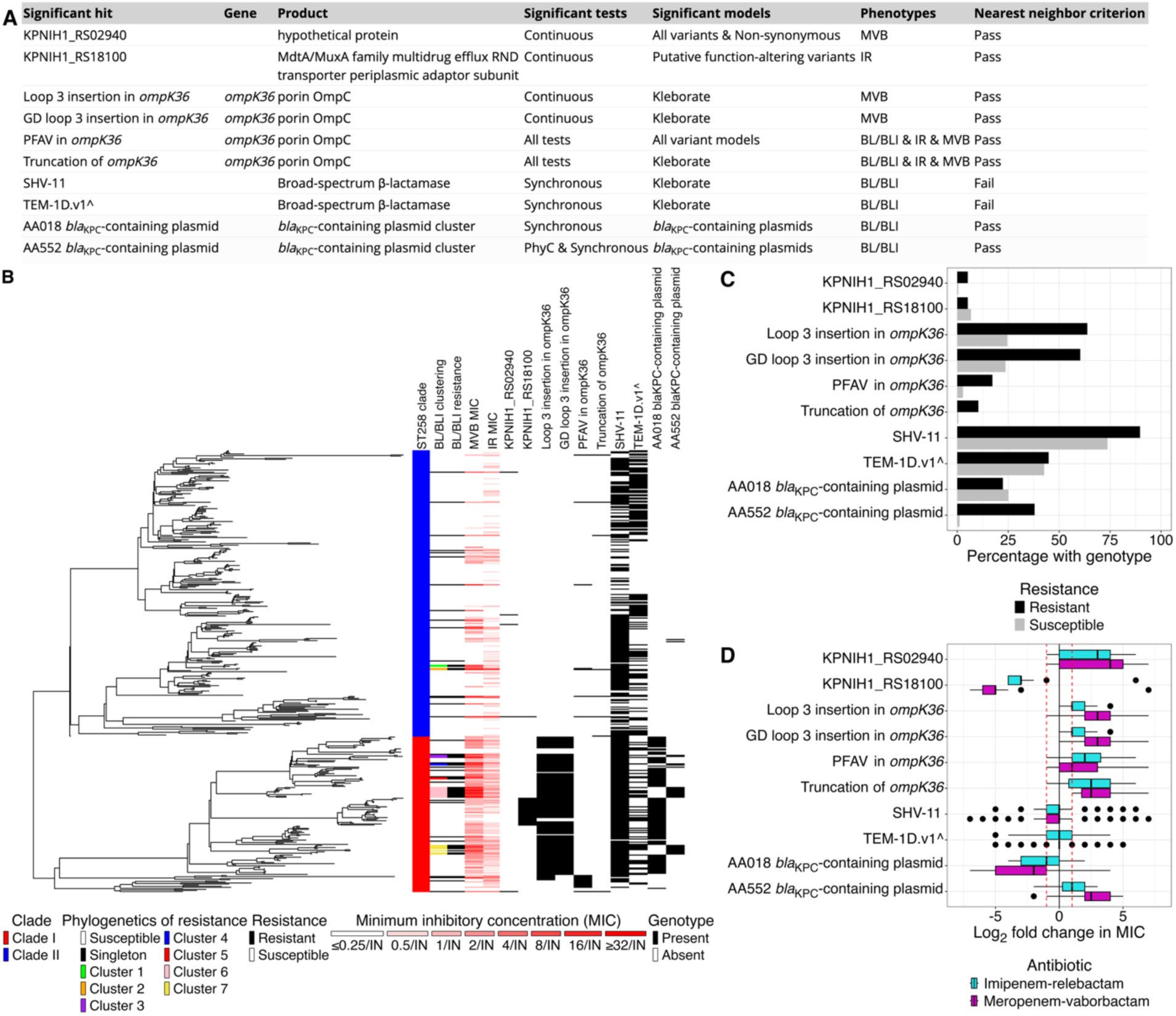
All significant genome-wide association study hits for β-lactam/β-lactamase inhibitor resistance Convergence-based genome-wide association study was performed on log_2_ minimum inhibitory concentration and resistance to β-lactam/β-lactamase inhibitor combinations using hogwash v.1.2.6. Hits that passed the nearest neighbor criterion (median log_2_ fold change in MIC > |1|) were eligible for downstream analysis. (**a**) Information about each significant genome-wide association study hit. (**b**) The significant hits are overlaid on the phylogenetic tree. **c** The frequency of genotypes in β-lactam/β-lactamase inhibitor combination susceptible and resistant isolates. (**d**) The log_2_ fold change in imipenem-relebactam and meropenem-vaborbactam minimum inhibitory concentration for isolates with the genotype relative to their nearest phylogenetic neighbor. **Abbreviations:** BL/BLI, β-lactam/β-lactamase inhibitor; IR, imipenem-relebactam; MVB, meropenem-vaborbactam; PFAV, putative function-altering variant.

**Supplementary Figure 5.**
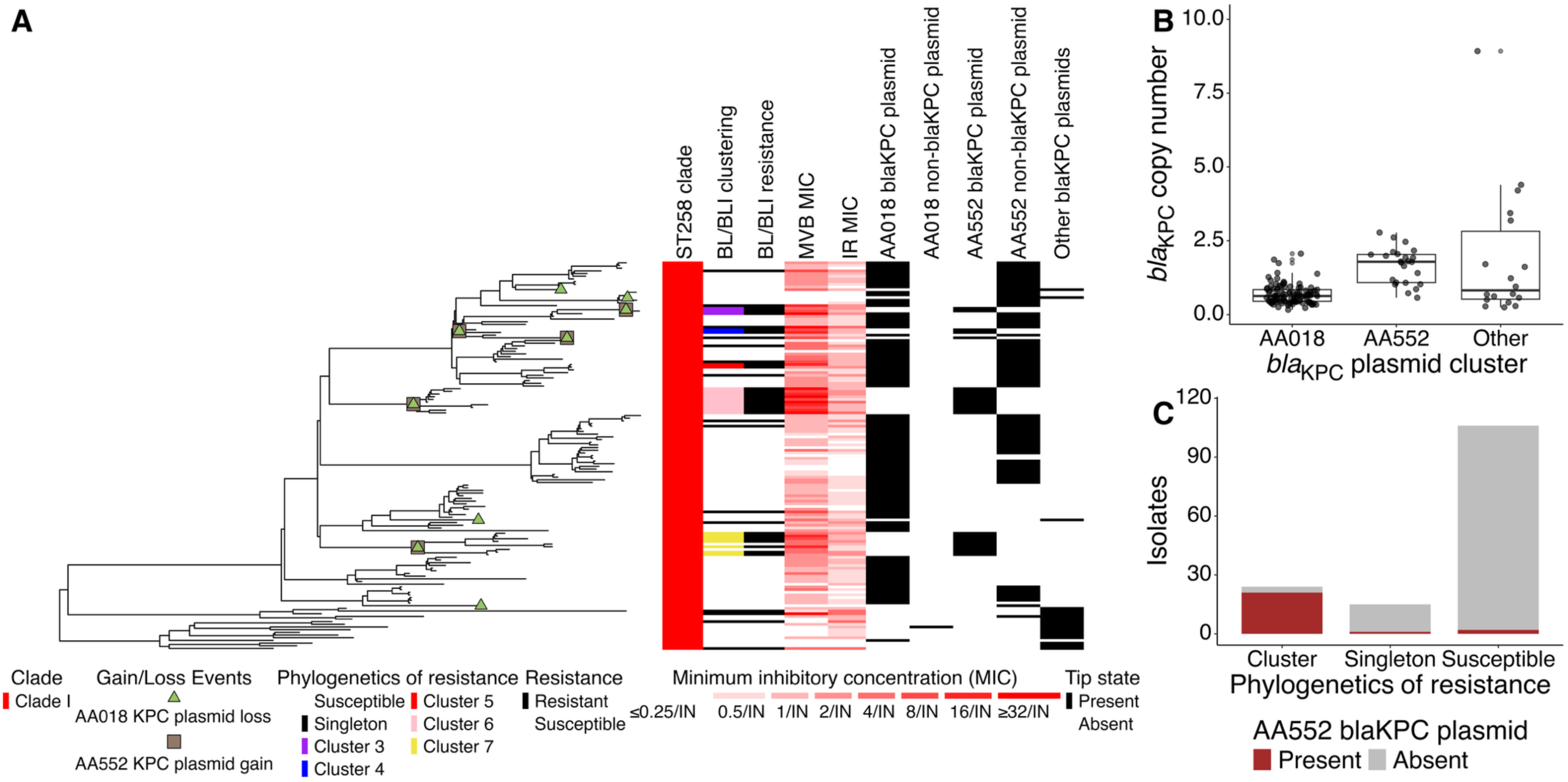
Differences in *bla*_KPC_ plasmid backbone contribute to elevated *bla*_KPC_ copy number and the spread of resistance in clade I (**a**) The presence of dominant *Klebsiella* pneumoniae carbapenemase (*bla*_KPC_)-containing plasmids across clade I. The emergence of resistance and select resistance-associated genotypes is overlaid on ancestral nodes. (**b**) Differences in *bla*_KPC_ copy number across *bla*_KPC_ plasmids. (**c**) The frequency of *bla*_KPC_-containing AA552 plasmids in β-lactam/β-lactamase inhibitor resistant clusters, singletons, and susceptible isolates. **Abbreviations:** BL/BLI, β-lactam/β-lactamase inhibitor; IR, imipenem-relebactam; MVB, meropenem-vaborbactam; PFAV, putative function-altering variant.

**Supplementary Figure 6.**
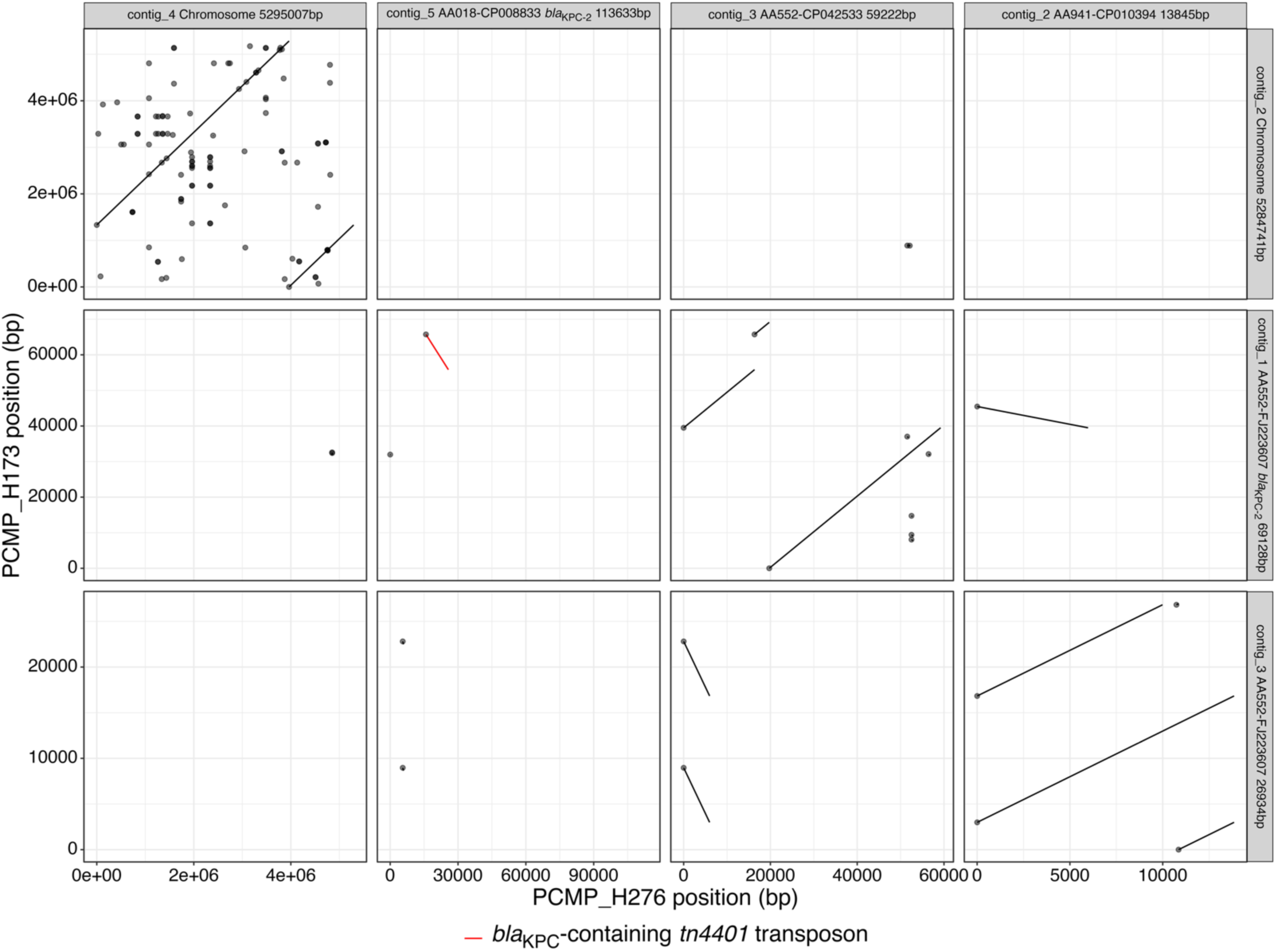
Evidence of the *tn4401 bla*_KPC_ transposon hopping from AA018 plasmid to the AA552 plasmid Long-read sequencing was performed on a representative clade I isolate with *Klebsiella pneumoniae* carbapenemase (*bla*_KPC_)-containing AA552 plasmid and a closely related isolate with a *bla*_KPC_-containing AA018 plasmid. Grids were named using the isolate’s contig name, Mob-Suite plasmid calls (primary cluster – mash nearest neighbor), the presence of the *bla*_KPC_ gene, and contig size. The presence of the bla_KPC_ transposon, *tn4401*, was labeled in red.

**Supplementary Figure 7.**
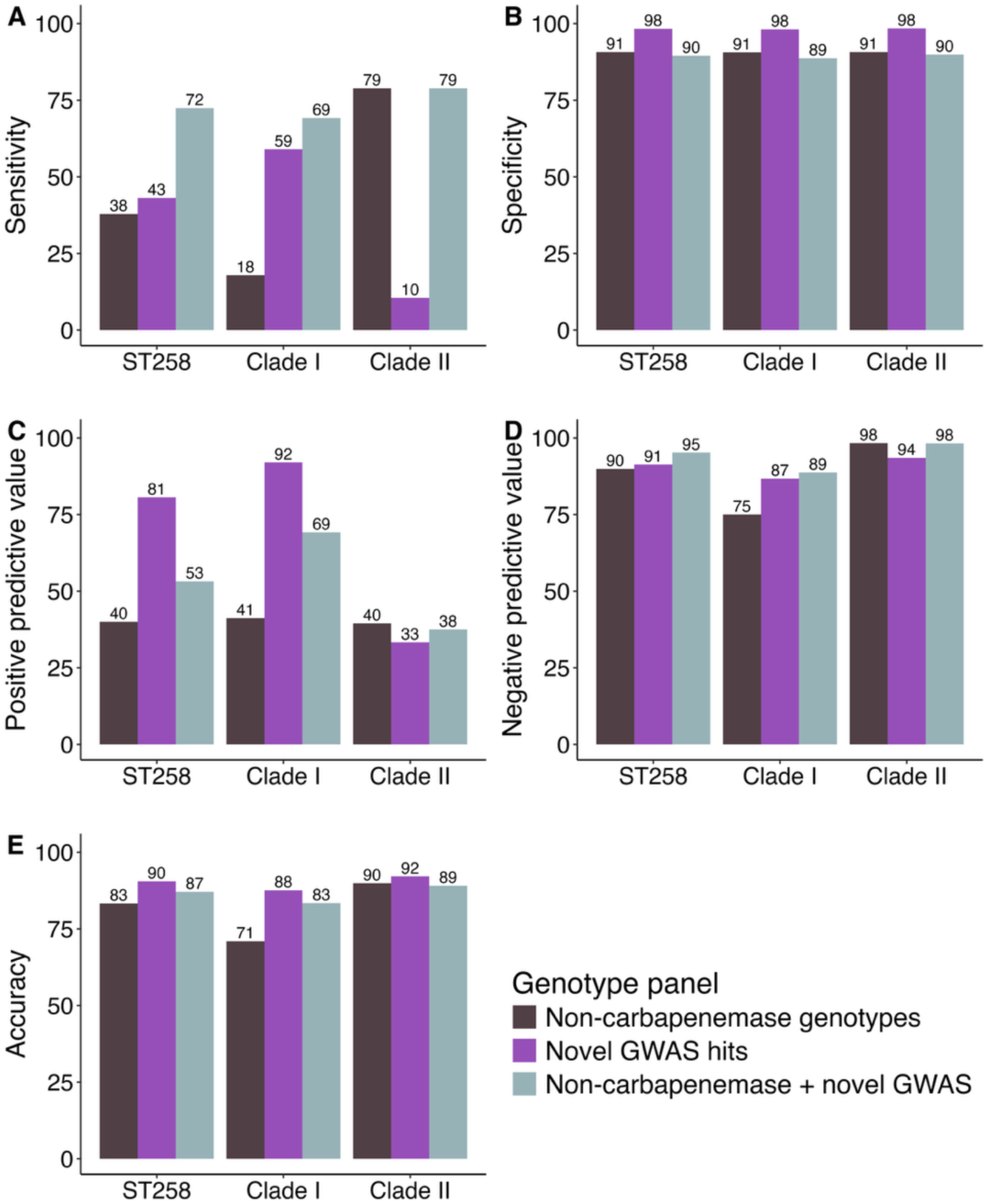
Performance statistics for resistance and our genotype panels The sensitivity (**a**), specificity (**b**), positive predictive value (**c**), negative predictive value (**d**), and accuracy (**e**) of our genotype panels for resistance to β-lactam/β-lactamase inhibitor combinations. The following genotypes were considered in our known carbapenem resistance-associated panel: truncations and putative function-altering variants in *ompK36*, insertion sequences near the *ompK36* promoter, putative function-altering variants in *ramA* efflux pump activator, and non-synonymous mutations in PBPs. The following novel hits were identified in our genome-wide association study: *Klebsiella pneumoniae* carbapenemase (*bla*_KPC_)-containing AA552 plasmid and the hypothetical protein, KPNIH1_RS02940. **Abbreviations:** BL/BLI, β-lactam/β-lactamase inhibitor; IR, imipenem-relebactam; MVB, meropenem-vaborbactam; PFAV, putative function-altering variant.

**Supplementary Figure 8.**
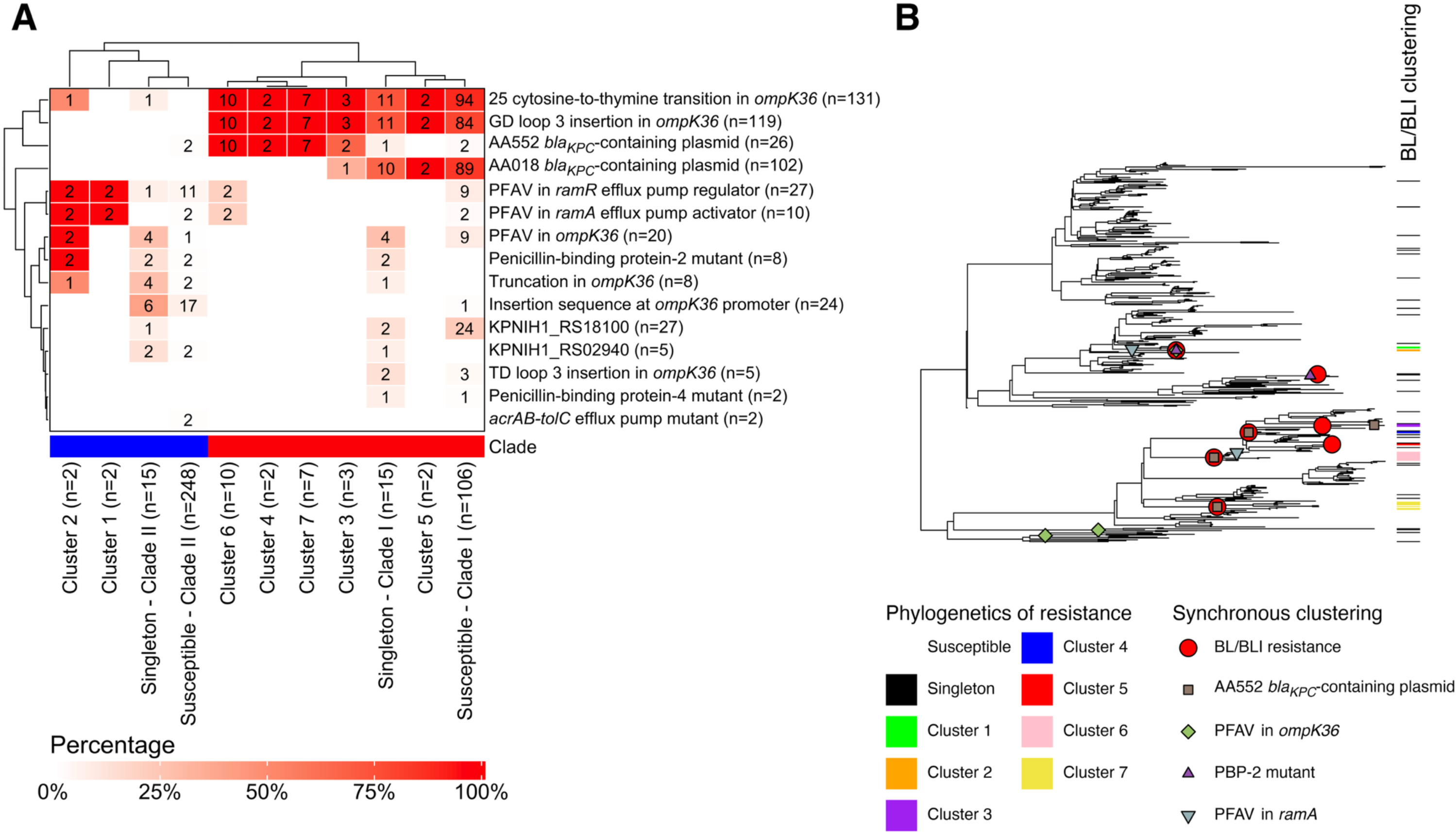
Differential potential for emergence and spread of β-lactam/β-lactamase inhibitor resistance-associated genotypes (**a**) The frequency of genotypes in phylogenetic singletons of β-lactam/β-lactamase inhibitor (BL/BLI) resistance, clusters of BL/BLI resistance, and susceptible isolates. (**b**). The synchronous clustering of BL/BLI resistance and resistance genotypes is overlaid on ancestral nodes. **Abbreviations:** BL/BLI, β-lactam/β-lactamase inhibitor; PFAV, putative function-altering variants; PBP, penicillin-binding protein.

**Supplementary Figure 9.**
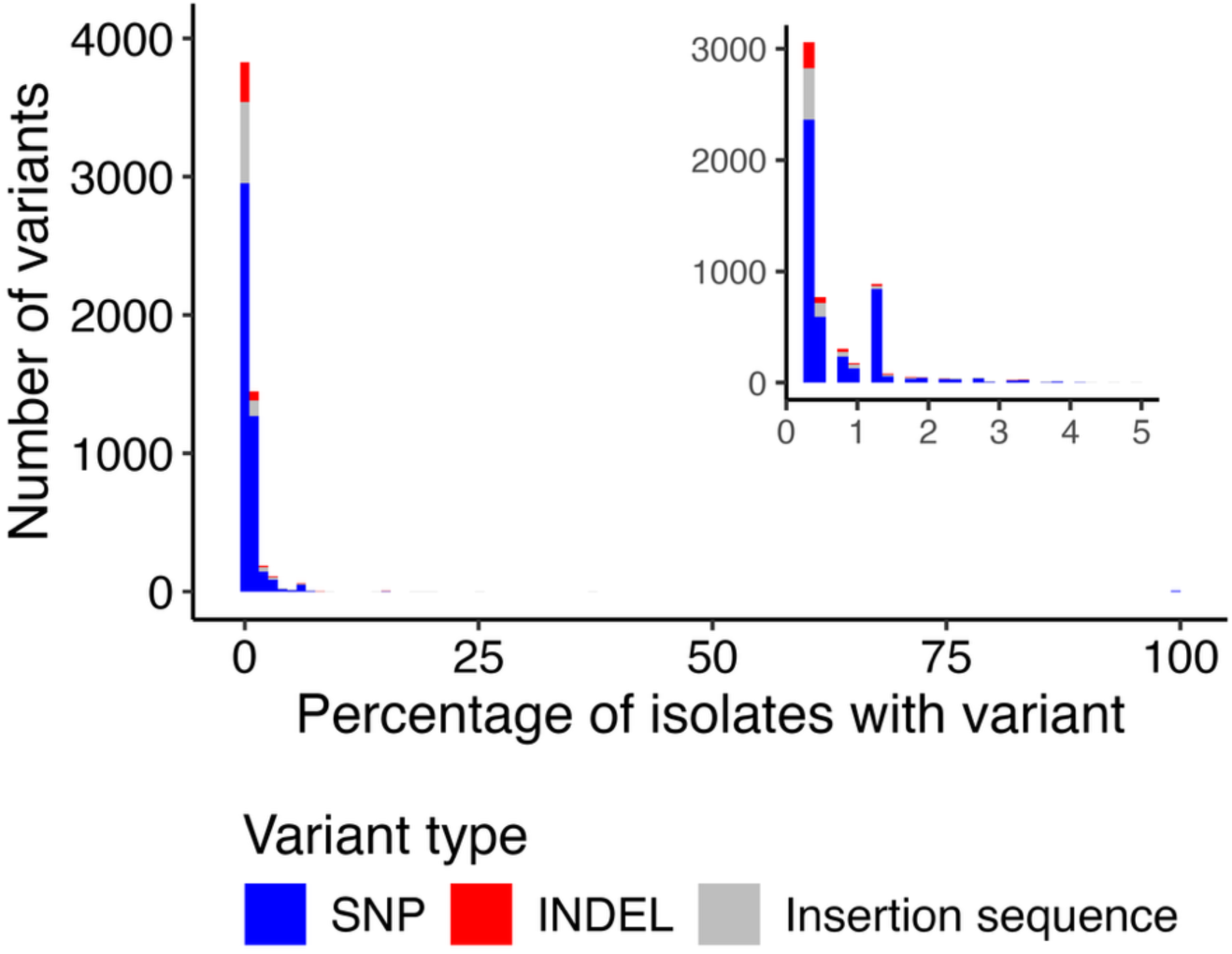
Percentage of isolates with a core genome variant The percentage of isolates with considered variants: single-nucleotide polymorphisms, insertions, deletions, and insertion sequences. The figure insert highlights the distribution of variants that were found in less than five percent. **Abbreviations:** INDEL, insertion or deletion; SNP, single-nucleotide polymorphism

## Notes

### Author Declarations

No new data used. The original study was reviewed and approved by the Institutional Review Board of the University of Pennsylvania with a waiver of informed consent.

